# Dermatomyositis: Muscle Pathology According to Antibody Subtypes

**DOI:** 10.1101/2021.06.03.21258156

**Authors:** Jantima Tanboon, Michio Inoue, Yoshihiko Saito, Shinichiro Hayashi, Satoru Noguchi, Naoko Okiyama, Manabu Fujimoto, Ichizo Nishino

## Abstract

**Importance:** Current pathological criteria of dermatomyositis (DM) do not recognize different features among DM subtypes classified by dermatomyositis-specific antibodies (DMSAs).

**Objective:** To determine whether myopathological features differ among DM subtypes classified by DMSAs and whether the pathological features can be characterized by serologically defined DM subtype.

**Design:** Retrospective review of muscle pathology slides of 256 patients diagnosed with DM from January 2009 to December 2020.

**Setting:** Single center study in a tertiary laboratory for muscle diseases.

**Participants:** A total of 256 patients whose DM diagnosis was pathologically confirmed based on the sarcoplasmic expression of myxovirus resistant protein A (MxA) were included. Of these, 249 patients were positive for one of the 5 DMSAs (seropositive patients, anti-TIF1-γ=87, anti-Mi-2=40, anti-MDA5=29, anti-NXP-2=83, and anti-SAE=10), and 7 were negative for all 5 DMSAs (seronegative patients).

**Exposure:** Histochemical, enzyme histochemical, immunohistochemical staining, and ultrastructural study.

**Main outcomes and measures:** Histological features stratified according to four pathology domains: muscle fiber, inflammatory, vascular, and connective tissue domains, and histological features of interest by histochemistry, enzyme histochemistry, and immunohistochemical study commonly used in the diagnosis of inflammatory myopathy.

**Results:** DMSAs significantly associated with characteristic histochemical and immunohistochemical features were as follows: anti-TIF1-γ with vacuolated/punched out fibers (64.7%, *P*<.001) and perifascicular enhancement in HLA-ABC (75.9%, *P*<.001); anti-Mi-2 with prominent muscle fiber damage (score 4.8±2.1, *P*<.001), inflammatory cell infiltration (score 8.0±3.0, *P*=.002), perifascicular atrophy (67.5%, *P*=.02), perifascicular necrosis (52.5%, *P*<.001), increased perimysium alkaline phosphatase activity (70.0%, *P*<.001), central necrotic peripheral regenerating fibers (45.0%, *P*<.001), and sarcolemmal deposition of the membrane attack complex (67.5%, *P*<.001); anti-MDA5 with scattered/diffuse staining pattern of MxA (65.5%, *P*<.001) with less muscle pathology and inflammatory features; and anti-NXP2 with microinfarction (26.5%, *P*<.001); and anti-SAE and seronegative DM with HLA-DR expression (50.0%, *P*=.02 and 57.1%, *P*=.02 respectively).

**Conclusion and relevance:** We described an extensive study on serological-pathological correlation of DM primarily using MxA expression as an inclusion criterion. DMSAs was associated with distinctive myopathological features in our studied cohort, suggesting that different pathobiological mechanisms may underscore each subtype.

**Key points:** *Question:* Are myopathological features different among dermatomyositis (DM) subtypes classified by DM-specific autoantibodies (DMSAs)? If so what are the characteristic features of each subtype?

*Findings:* This study enrolled 256 (249 DMSA-positive and 7 seronegative) patients whose DM diagnosis was made pathologically by confirming the expression of myxovirus resistant protein A in the sarcoplasm of muscle fibers in muscle biopsy samples. The DM subtypes classified by the positive DMSAs were associated with distinctively characteristic pathological features.

*Meaning:* Different pathological features suggest different pathological mechanisms may well underly each DM subtype classified by DMSA.

## Introduction

The diagnostic criteria of dermatomyositis (DM) have transformed over the past four decades from clinically oriented criteria, primarily based on the presence of skin lesions and muscle weakness described by Bohan and Peter in 1975, to the clinical-serological-pathological criteria proposed by the European Neuromuscular Center (ENMC) in 2018 (2018 ENMC-DM).^1-3^ In 2018 ENMC-DM, the following 5 DM-specific antibodies (DMSAs) were included as the serological criteria: anti-transcription intermediary factor 1-γ (TIF1-γ), anti-complex nucleosome remodeling histone deacetylase (Mi-2), anti-melanoma differentiation gene 5 (MDA5), anti-nuclear matrix protein 2 (NXP-2), and anti-small ubiquitin-like modifier-activating enzyme (SAE).^2^ In addition to perifascicular atrophy (PFA), the best known pathological feature of DM, the 2018 ENMC-DM included myofiber expression of myxovirus resistant protein A (MxA), the surrogate marker for type I interferon (IFN1) pathway activation, as a diagnostic finding.^4-7^ Interestingly, DMSA-associated clinical phenotypes have been characterized, including anti-TIF1-γ DM with DM skin lesions, dysphagia, and malignancy;^8-10^ anti-Mi-2 DM with high serum creatine kinase (CK) level, myalgia, and muscle weakness; ^11-13^ anti-MDA5 DM with mechanic hands and interstitial lung disease (ILD), but low CK levels and less muscle involvement;^14, 15^ and anti-NXP-2 DM with muscle weakness, but less skin involvement.^16,17^ Conversely, DMSA-associated pathological phenotypes have been limited to small studies. ^8, 11-13,18,19^ This study aimed to investigated and characterize DMSA-specific pathological features.

## Methods

### Patients

Muscle biopsies from 256 patients pathologically diagnosed with DM at the National Center of Neurology and Psychiatry (NCNP), a nationwide referral center for muscle disease in Japan, from January 2009 to December 2020 were evaluated by confirming the expression of MxA in the sarcoplasm of the muscle fibers, which are neither necrotic nor regenerating.^5-7,20^ Because of the existence of DM *sine* dermatitis (DMSD),^16^we regarded all MxA-positive muscle biopsies as DM regardless of the presence of skin lesion. This study was an expansion of the DM cohort from our previous studies^11,16^ and consisted of 249 muscle biopsies from patients positive for one of the 5 DMSAs and 7 muscle biopsies from patients who were tested but negative for all the 5 DMSAs (seronegative DM). We defined patients <18 years as juvenile patients.

### Serological information

ELISA for autoantibodies against TIF1-γ, Mi-2, and MDA5 were covered by the national health insurance system in Japan (MESACUP™ kit, Medical & Biological Laboratories Co, Ltd, Nagoya, Japan) and were performed according to the manufacturer’s instructions. Patients testing negative for anti-TIF1-γ, Mi-2, and MDA5 antibodies were evaluated by immunoprecipitation and western blotting for autoantibodies against NXP-2 and SAE.^21-23^

### Histochemical and immunohistochemical evaluation

Histochemical and immunohistochemical staining was performed for routine diagnostic purposes. Details regarding immunohistochemical stains are available in the Supplement. JT performed pathological evaluation and was blinded to the antibody results at the time of evaluation. The histochemical and immunohistochemical stained slides, prepared at the time of pathological diagnosis at the NCNP, were evaluated based on 4 domains (muscle fiber, inflammatory, vascular, and connective tissue domains) using the same scoring system as our previous studies modified from the pathology scoring system originally used for juvenile DM (supplement and eTable 1).^11, 24^ Alongside the PFA, perifascicular necrosis (PFN), decreased cytochrome c oxidase (COX) activity in perifascicular area, perivascular vascular inflammation, vasculitis, CD8 and acid phosphatase (ACP)/CD68 infiltration in non-necrotic fiber, and CD20 aggregation, we included microinfarction, centrally necrotic-and-peripherally regenerating (CNPR) fibers, and vacuolated/punched-out fiber as histopathology of interest for DM. Microinfarction was defined by the aggregate of at least 3 necrotic fibers without inflammatory cell infiltration into the necrotic fiber accompanied by decreased/absent sarcoplasmic oxidative enzymatic activity on nicotinamide adenine dinucleotide dehydrogenase-tetrazolium reductase. CNPR fibers were defined as fibers presenting necrosis in the central part of the sarcoplasm surrounded by crescent-shaped regenerating portions. Vacuolated/punched out fibers were defined by degenerative non-necrotic fiber with vacuolations.^8^ For MxA staining, 3 patterns were documented: 1) perifascicular pattern if the sarcoplasmic staining limited to the PFA; 2) scattered/diffuse pattern if there was no specific localization; and 3) mixed pattern. The evaluation criteria for histochemical and immunohistochemical staining are described elsewhere.^11^ Ultrastructural evaluation for tubuloreticular inclusions (TRIs) was performed in 54 patients.

## Statistical analysis

We explored histological features by performing multiple correspondence analysis (MCA) followed by agglomerative hierarchical clustering (AHC) using XLSTAT version 2021.2.1 New Yok, USA, www.XLSTAT.com. For continuous variables, Welch’s one-way ANOVA followed by Dunnett’s T3 multiple comparisons test was performed. Welch’s t-test and the Fisher exact test were used for continuous and categorial variables to compare findings in one antibody subtype against other antibody subtypes (e.g. anti-TIF1-γ DM vs. non-TIF1-γ DM); the tests were 2 tailed. A *P*-value <0.05 was regarded as statistically significant. These analyses were performed using GraphPad Prism version 9.1.0 (216) for Mac OS, GraphPad Software, San Diego, California USA).

### Standard protocol approvals, registrations, and patient consent

This study was approved by the institutional review boards of the NCNP. All clinical information and materials derived from diagnostic testing and was permitted for research use with written informed consent from the patients.

## Results

### Patient characteristics

Of the 256 patients pathologically diagnosed with DM by sarcoplasmic MxA expression, 249 were DMSA-positive (n=249, anti-TIF1-γ=87, anti-Mi-2=40, anti-MDA5=29, anti-NXP-2=83, anti-SAE=10) and 7 were seronegative. Clinical information of DM patients is summarized in Table 1. There were significant differences in patient ages among subgroups classified by positive DMSA (*P*<.001) (Figure 1A). The patients testing positive for anti-TIF1-γ, anti-Mi-2, and anti-SAE antibodies were older than non-TIF1-γ DM (42.3±26.2 years old, *P*<.001), non-Mi-2 DM (45.3±27.6 years old, *P*=.03), and non-SAE DM (45.6±6.8 years old, *P*<.001), respectively. Patients with anti-TIF1-γ and anti-Mi-2 DM were significantly younger than patients with anti-SAE DM (*P*=.003 and *P*=.002, respectively, Figure 1A). Anti-NXP-2 and seronegative DM affected younger individuals (non-NXP-2 DM 51.8± 24.9 years old, *P*<.001 and seropositive DM 47.4±26.4 years old, *P*=.007, respectively). This was reflected by the larger number of juvenile patients with anti-NXP-2 (42.2% vs. non-NXP-2 18.5%, *P*<.001) and seronegative DM (85.7% vs. seropositive 24.5%, *P*=.002). Disease duration, the proportions of patients receiving systemic immunotherapy within 6 weeks before the time of muscle biopsy, the sex of affected patients, and the biopsy site, did not significantly differ among DMSA subtypes (eTable 2). In anti-TIF1-γ DM, biopsies obtained from biceps brachii (42.5%) were slightly less common than non-TIF1-γ DM (52.5%, *P*=.05). The CK levels were distinctively different among DMSA subtypes (*P*<.001) (Figure 1B). Anti-Mi-2 DM were associated with higher CK levels (8113.7±14314.2 vs. non-Mi-2 2758.2±5566.7 U/L, *P*=.03), while anti-MDA5, anti-SAE, and anti-TIF1-γ DM were associated with lower CK levels (394.6±418.9 vs. non-MDA5 4009.3±8232.0 U/L, *P*<.001; 492.8±357.0 vs. non-SAE 3725.0±7967.2 U/L, *P*<.001; and 1881.2±4414.3 vs. non-TIF1-γ 4472.0±8981.1 U/L, *P*=0.002, respectively). Anti-TIF1-γ DM patients more commonly presented with skin lesions of any kind (97.7% vs. non-TIF1-γ 87.0% DM, *P*=.006) as well as DM specific skin lesions defined by the 2018 ENMC-DM criteria (89.7% vs. non-TIF1-γ 60.4%, *P*<.001). They were also significantly associated with dysphagia (47.1% vs. non-TIF1-γ 28.0%, *P*=.003) and malignancy (41.4% vs. non-TIF1-γ 11.2%, *P*<.001). However, for anti-TIF1-γ DM patients under age 50 years, the association with malignancy (11.1%) compared to non-TIF1-γ DM was not statistically significant (5.3%, *P*=0.65). Anti-Mi-2 DM patients more frequently presented with muscle weakness (100% vs. non-Mi-2 87.5%, *P*=.01). Anti-MDA5 DM was significantly associated with mechanic’s hands (51.7% vs. non-MDA5 13.2%, *P*<.001) and ILD (79.3% vs. non-MDA5 10.6%, *P*<.001), but were significantly less associated with muscle weakness (69.0% vs. non-MDA5 92.1%, *P*=.001) and malignancy (3.5% vs. non-MDA5 23.8%, *P*=.008). Anti-NXP-2 DM was less likely to be associated with skin lesion of any kind (77.1% vs. non-NXP-2 97.1%, *P*<.001), DM specific skin lesions (44.6% vs. non-NXP-2 82.7%, *P*<.001), mechanic’s hands (3.6% vs. non-NXP-2 22.3%, *P*<.001), ILD (8.4% vs. non-NXP-2 23.3%, *P*=.004) and malignancy (8.4% vs. non-NXP-2 27.8%, *P*<.001). Myalgia was distinctively more common in anti-NXP-2 DM (84.2%) than non-NXP-2 DM (61.6%, *P*<.001). Anti-SAE was not associated with any distinctive clinical feature. Seronegative DM was less associated with typical DM skin lesions (28.6%) than seropositive DM (71.5% *P*=.03).

**Table 1.** Clinical Features of all Dermatomyositis Cases in this Study.

|  | All DM | Anti-TIF1-γ | Anti-Mi-2 | Anti-MDA5 | Anti-NXP-2 | Anti-SAE | Sero-negative |
| --- | --- | --- | --- | --- | --- | --- | --- |
|  | (n=256) | (n=87) | (n=40) | (n=29) | (n=83) | (n=10) | (n=7) |
| **Clinical features** |  |  |  |  |  |  |  |
| Age at biopsy, y | 46.5 ± 26.7 | 54.8 ± 25.8* | 53.2 ± 20.0* | 42.5 ± 23.7 | 35.6 ± 27.1* | 70.4 ± 8.1* | 17.3 ± 19.9* |
| Juvenile (<18 years) | 67 (26.2) | 15 (17.2)* | 3 (7.5)* | 8 (27.6) | 35(42.2)* | 0 | 6 (85.7)* |
| Sex, female | 139(54.3) | 48 (55.2) | 18 (45.0) | 16 (55.2) | 47 (55.6) | 7 (70.0) | 3 (42.9) |
| CK (U/L) | 3598.2 ± 7834.3<sup>a</sup> | 1881.2 ± 4414.3<sup>a,*</sup> | 8113.7 ± 14314.2* | 394.6 ± 481.9* | 4685.2 ± 7035.0 | 492.8 ± 357.0* | 3710.4 ± 7248 |
| Skin lesions<sup>b</sup> | 232 (90.6) | 85 (97.7)* | 37 (92.5) | 29 (100.0) | 64 (77.1)* | 10 (100.0) | 7(100.0) |
| DM skin lesion(s)<sup>c</sup> | 180 (70.3) | 78 (89.7)* | 29 (72.5) | 25 (86.2) | 37 (44.6)* | 9 (90.0) | 2 (28.6)* |
| Mechanic hands | 45 (17.6) | 16 (18.4) | 8(20.0) | 15 (51.7)* | 3 (3.6)* | 3 (30.0) | 0 |
| Muscle weakness | 229 (89.5) | 75 (86.2) | 40 (100.0)* | 20 (69.0)* | 79 (95.2)* | 9 (90.0) | 6 (85.7) |
| Myalgia | 175(68.9)<sup>d</sup> | 59 (67.8) | 23 (57.5) | 15(53.6)<sup>a</sup> | 69 (84.1)<sup>a,*</sup> | 4 (40.0) | 5 (71.4) |
| Dysphagia | 88 (34.5)<sup>a</sup> | 41 (47.1)* | 8 (20.5)<sup>a</sup> | 8 (27.6) | 25(30.1) | 6 (60.0) | 0 |
| Interstitial lung disease | 47 (18.4)<sup>e</sup> | 11 (12.6) | 3 (7.7)<sup>e</sup> | 23 (79.3)* | 7 (8.4)* | 3 (30.0) | 0 |
| Malignancy | 55(21.5) | 36 (41.4)* | 7 (17.5) | 1 (3.4)* | 7(8.4)* | 4 (40.0) | 0 |
| Malignancy, age <50 years | 5(2.0) | 4(4.6) | 0 | 0 | 1(1.2) | 0 | 0 |

**Abbreviations:** DM, dermatomyositis; anti-TIF1-γ, anti-transcription intermediary factor 1-gamma antibody; anti-Mi-2, anti-complex nucleosome remodeling histone deacetylase antibody; anti-MDA5, anti-melanoma differentiation-associated gene 5; anti-NXP-2, anti-nuclear matrix protein 2; anti-SAE, anti-small ubiquitin-like modifier activating enzyme; CK, creatine kinase.
Continuous data are shown as mean ± SD; categorical data are shown as number (percentage).
<sup>a</sup>Information was not available for 1 patient
<sup>b</sup>Any type of skin lesion
<sup>c</sup>DM skin lesions as defined by the 239th European Neuromuscular Centre workshop: presence of Gottron sign/papule and/or heliotrope rash
<sup>d</sup>Information was not available for 2 patients
\* $p < 0.05$ (vs. other antibody subtypes, red-positive association, blue-negative association)

**Figure 1.**
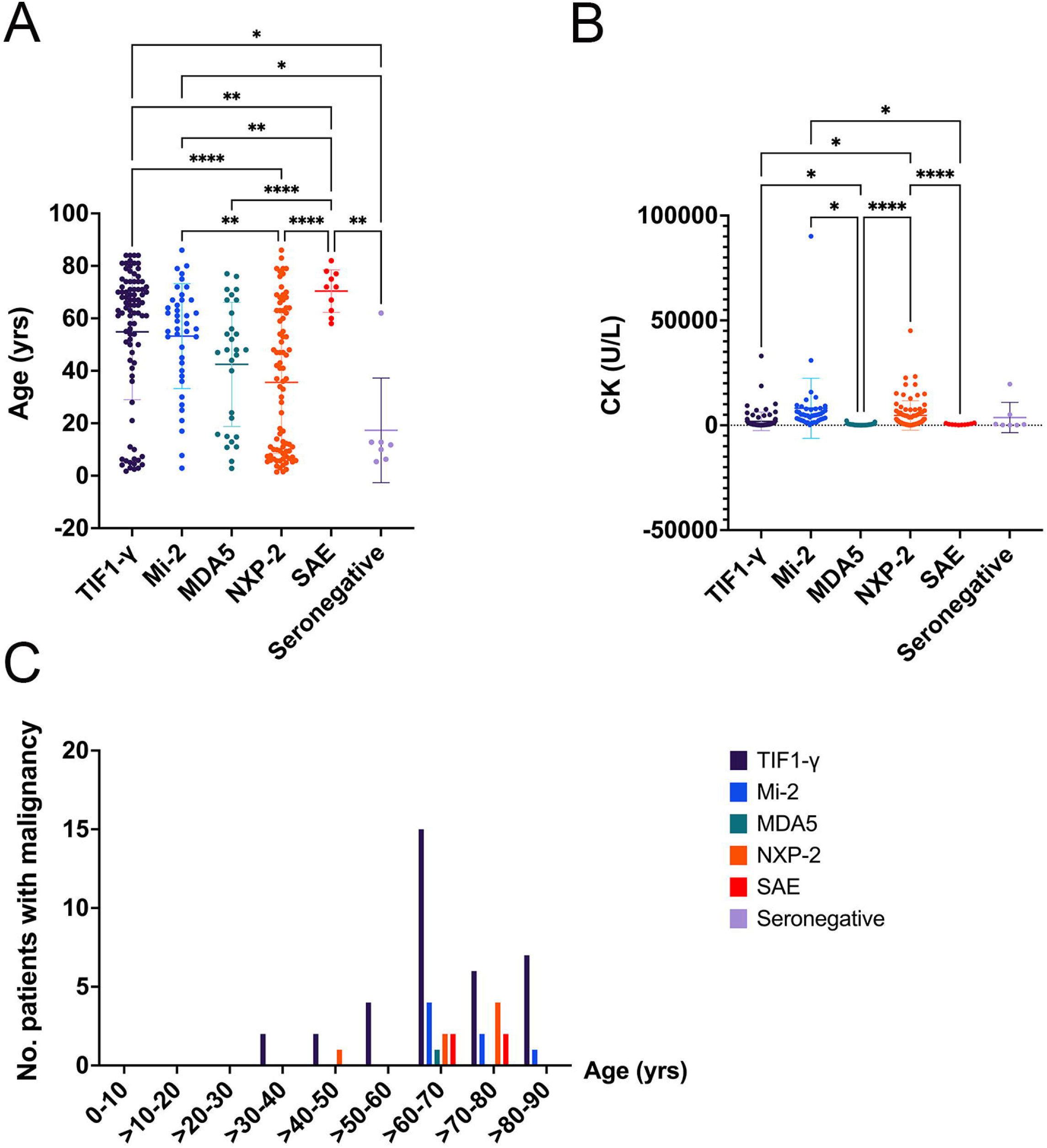
Age distribution, CK level, and malignancy association among DMSA subtypes in this study. **A. Age at biopsy of dermatomyositis (DM) patients in this study**. Although most patients (74.8%) were adults, a significantly higher proportion of juvenile patients presented anti-NXP-2 antibodies (42.2%) and seronegative DM (85.7%). Age at biopsy of anti-NXP-2 DM patients was significantly lower than the age of anti-TIF1-γ, anti-Mi-2, and anti-SAE DM patients. **B. CK level at the time of muscle biopsy**. Anti-MDA5 DM was associated with low CK level; the level was significantly lower than anti-TIF1-γ, anti-Mi-2, and anti-NXP-2 DM. **C. Malignancy-associated dermatomyositis**. Anti-TIF1-γ DM was significantly associated with malignancy (36/87, 41.4%) and was associated with the highest proportion of malignant patients under age 50 years (4/87, 4.6%). **Note:** bar=mean±SD, ANOVA with Dunnette’s T3 multiple comparison test ^*^p<0.0332, ^**^p<0.0021, ^***^p<0.0002, ^****^p<0.0001 **Abbreviations:** CK, creatine kinase; DM, dermatomyositis; DMSA, dermatomyositis-specific antibody; yrs, years.

### Ultrastructural study

TRIs were observed in all 54 muscle biopsies (anti-TIF1-γ=17, anti-Mi-2=7, anti-MDA5=5, anti-NXP-2=22, anti-SAE=1, seronegative=2).

### Histological features and clustering

We used 6 antibody subtypes (5 DMSAs and 1 seronegative) and 23 histologic variables for exploratory MCA and AHC analyses (Table 2 and eTable 3). We proposed 6 pathology classes by truncation. Pathological features in anti-Mi-2, anti-MDA5, and anti-SAE and seronegative DM were more distinctive as classes 1, 3, and 6 were composed of 74.4% of anti-Mi-2, 72.4% of anti-MDA5, and 90.0% of anti-SAE and 100% of seronegative DM, respectively. The features in anti-TIF1-γ and anti-NXP-2 DM were more similar (eTable 3).

**Table 2.** Histological and Immunohistological Features of Dermatomyositis in this Study.

|  | All DM | Anti-TIF1-γ | Anti-Mi-2 | Anti-MDA5 | Anti-NXP-2 | Anti-SAE | Sero-negative |
| --- | --- | --- | --- | --- | --- | --- | --- |
|  | (n=256) | (n=87) | (n=40) | (n=29) | (n=83) | (n=10) | (n=7) |
| **Muscle fiber domain** |  |  |  |  |  |  |  |
| Muscle fiber domain score | 2.9 ± 2.1 | 2.9 ± 1.9 | 4.9 ± 2.1* | 1.1 ± 1.6* | 2.6 ± 2.0 | 3.8 ± 2.2 | 2.9 ± 1.2 |
| Necrotic fiber score | 0.9 ± 0.9 | 0.8 ± 0.8 | 1.8 ± 0.5* | 0.3 ± 0.6* | 0.9 ± 0.9 | 1.0 ± 0.8 | 0.4 ± 0.5* |
| Regenerating fiber score | 0.5 ± 0.5 | 0.4 ± 0.5* | 0.8 ± 0.4* | 0.2 ± 0.4* | 0.5 ± 0.5 | 0.6 ± 0.5 | 0.9 ± 0.4* |
| Atrophic fiber score | 0.5 ± 0.8 | 0.5 ± 0.7 | 0.9 ± 0.9* | 0.2 ± 0.6* | 0.4 ± 0.6* | 0.7 ± 0.8 | 0.3 ± 0.8 |
| PFA score | 0.8 ± 0.9 | 0.9 ± 0.9 | 1.2 ± 0.9* | 0.3 ± 0.7* | 0.7 ± 0.9 | 1.3 ± 0.9 | 1.0 ± 1.0 |
| Fiber with internalized nuclei>3% score | 0.2 ± 0.4 | 0.2 ± 0.4* | 0.2 ± 0.4 | 0.0 ± 0.2* | 0.1 ± 0.3 | 0.2 ± 0.4 | 0.3 ± 0.5 |
| **Inflammatory domain** |  |  |  |  |  |  |  |
| Inflammatory domain score | 6.6 ± 3.3<sup>a</sup> | 6.6 ± 3.4 | 8.0 ± 3.0<sup>a</sup>* | 3.7 ± 2.4* | 6.8 ± 2.9 | 5.9 ± 3.3 | 8.9 ± 3.8 |
| Endomysial CD3 infiltration score | 1.0 ± 0.7<sup>a</sup> | 1.0 ± 0.7 | 1.3 ± 0.6<sup>a</sup>* | 0.6 ± 0.6* | 1.0 ± 0.6 | 0.9 ± 0.6 | 1.6 ± 0.8 |
| Perimysial CD3 infiltration score | 0.5 ± 0.7<sup>a</sup> | 0.6 ± 0.7 | 0.7 ± 0.7<sup>a</sup> | 0.1 ± 0.4* | 0.5 ± 0.7 | 0.6 ± 0.8 | 1.0 ± 1.0 |
| Endomysial CD20 infiltration score | 1.0 ± 0.8<sup>a</sup> | 1.0 ± 0.7 | 1.5 ± 0.7<sup>a</sup>* | 0.3 ± 0.6* | 1.0 ± 0.7 | 0.6 ± 0.7 | 1.1 ± 1.0 |
| Perimysial CD20 infiltration score | 0.5 ± 0.7<sup>a</sup> | 0.6 ± 0.7 | 0.6 ± 0.7<sup>a</sup> | 0.1 ± 0.4* | 0.5 ± 0.7 | 0.4 ± 0.8 | 0.9 ± 0.7 |
| Endomysial CD68 infiltration score | 1.8 ± 0.4<sup>a</sup> | 1.8 ± 0.5 | 1.9 ± 0.3<sup>a</sup> | 1.6 ± 0.5* | 1.8 ± 0.4 | 1.8 ± 0.4 | 1.9 ± 0.3 |
| Perimysial CD68 infiltration score | 1.2 ± 0.7<sup>a</sup> | 1.2 ± 0.8 | 1.5 ± 0.6<sup>a</sup>* | 0.8 ± 0.7* | 1.3 ± 0.7 | 1.4 ± 0.5 | 1.6 ± 0.5 |
| Perivascular inflammatory cell infiltration score | 0.4 ± 0.5 | 0.5 ± 0.5 | 0.5 ± 0.5 | 0.1 ± 0.4* | 0.5 ± 0.5 | 0.2 ± 0.4 | 0.9 ± 0.4* |
| **Vascular domain** |  |  |  |  |  |  |  |
| Capillary: fiber ratio, adult patient | 0.7 ± 0.3† | 0.7 ± 0.3† | 0.8 ± 0.3† | 0.8 ± 0.3† | 0.7 ± 0.3† | 0.7 ± 0.3† | 0.9 |
| Capillary: fiber ratio, juvenile patient | 0.5 ± 0.3<sup>b</sup> | 0.5 ± 0.2<sup>b</sup> | 0.4 ± 0.1 | 0.8 ± 0.4 | 0.4 ± 0.2* | NA | 0.6 ± 0.3 |
| **Connective tissue domain** |  |  |  |  |  |  |  |
| Perimysium fragmentation | 182 (71.9)<sup>c</sup> | 61(71.8)<sup>d</sup> | 33 (84.6)<sup>b</sup> | 11 (37.9)* | 63 (75.9) | 7 (70.0) | 7 (100.0) |
| Perimysium ALP activity, increased | 92 (35.9) | 21 (24.1)* | 28 (70.0)* | 4 (13.8)* | 33 (39.8) | 2 (20.0) | 4 (57.1) |
| Endomysial fibrosis | 37 (14.6)<sup>c</sup> | 12 (14.1)<sup>d</sup> | 9 (23.1)<sup>b</sup> | 1 (3.4) | 10 (12.0) | 1 (10.0) | 4 (57.1)<sup>*</sup> |
| **Histologic features of interest** |  |  |  |  |  |  |  |
| Perifascicular atrophy | 127(49.6) | 49(56.3) | 27(67.5)<sup>*</sup> | 7(24.1)<sup>*</sup> | 33(39.8)<sup>*</sup> | 7(70.0) | 4(57.1) |
| Perifascicular necrosis | 28(10.9) | 3(3.4)<sup>*</sup> | 21(52.5)<sup>*</sup> | 0 | 2(2.4)<sup>*</sup> | 0 | 2(28.6) |
| Decreased COX activity in perifascicular area | 118 (46.3)<sup>b</sup> | 47(54.7)<sup>b</sup> | 21(52.5) | 5(17.2)<sup>*</sup> | 37(44.6) | 4(40.0) | 4(57.1) |
| Perivascular inflammatory cell infiltration | 113(44.1) | 41(47.1) | 18(45.0) | 4(13.8)<sup>*</sup> | 42(50.6) | 2(20.0) | 6(85.7)<sup>*</sup> |
| Vasculitis | 39 (15.2) | 10 (11.5) | 10 (25.0) | 0 | 15 (18.1) | 0 | 4 (57.1)<sup>*</sup> |
| CD8 infiltration in non-necrotic fiber | 5 (2.0)<sup>a</sup> | 2 (2.3) | 3 (7.7)<sup>a,*</sup> | 0 | 0 | 0 | 0 |
| ACP/CD68 infiltration in non-necrotic fiber | 14 (5.5) | 4 (4.6) | 8 (20.0)<sup>*</sup> | 0 | 1 (1.2)<sup>*</sup> | 0 | 1 (14.3) |
| CD20 aggregation | 39 (15.3)<sup>a</sup> | 10 (11.5) | 11 (28.2)<sup>a,*</sup> | 1 (3.5) | 13 (15.7) | 1 (10.0) | 3 (42.9) |
| Microinfarction | 38 (14.8) | 11 (12.6) | 0 | 3 (10.3) | 22 (26.5)<sup>*</sup> | 2 (20.0) | 0 |
| Microinfarction, adult patients | 23 (12.2) | 10 (13.9) | 0 | 1 (4.8) | 10(20.8)<sup>*</sup> | 2 (20.0) | 0 |
| Microinfarction, juvenile patient | 15 (22.4) | 1 (6.7) | 0 | 2 (25.0) | 12(34.3)<sup>*</sup> | 0 | 0 |
| Central necrotic -peripheral regenerating fibers (CNPR) | 63 (24.6) | 21 (24.1) | 18 (45.0)<sup>*</sup> | 3 (10.3) | 18 (21.7) | 2 (20.0) | 1 (14.3) |
| Vacuolated, “punched out” fiber | 113 (44.5)<sup>d</sup> | 55(64.7)<sup>d,*</sup> | 3(7.5)<sup>*</sup> | 6(20.7)<sup>*</sup> | 40(48.2) | 7(70.0) | 2(28.6) |
| **Immunohistochemical features** |  |  |  |  |  |  |  |
| **MXA positivity** | 256(100.0) | 87(100.0) | 40(100.0) | 29(100.0) | 83(100.0) | 10(100.0) | 7(100.0) |
| MXA, perifascicular pattern | 96 (37.5) | 35 (40.2) | 27(67.5)<sup>*</sup> | 5(17.2)<sup>*</sup> | 23(27.7) | 3 (30.0) | 3 (42.9) |
| MXA, scattered/diffuse | 65 (25.4) | 14 (16.1)<sup>*</sup> | 5 (12.5)<sup>*</sup> | 19 (65.5)<sup>*</sup> | 24 (28.9) | 0 | 3 (42.9) |
| MXA, mixed pattern | 95 (37.1) | 38(43.7) | 8 (20.0)<sup>*</sup> | 5 (17.2)<sup>*</sup> | 36 (43.4) | 7 (70.0)<sup>*</sup> | 1 (14.3) |
| **HLA-ABC positivity** | 256(100.0) | 87(100.0) | 40(100.0) | 29(100.0) | 83(100.0) | 10(100.0) | 7(100.0) |
| HLA-ABC, perifascicular enhancement | 149 (58.2) | 66 (75.9)<sup>*</sup> | 26 (65.0) | 5 (17.2)<sup>*</sup> | 41 (49.4) | 8 (80) | 3 (42.9) |
| **HLA-DR positivity** | 46 (18.0) | 16 (18.4) | 5 (12.5) | 1 (3.4)<sup>*</sup> | 15 (18.1) | 5 (50.0)<sup>*</sup> | 4 (57.1)<sup>*</sup> |
| HLA-DR, perifascicular pattern | 27 (10.5) | 8 (9.2) | 1(2.5) | 0 | 10 (12.0) | 5 (50.0)<sup>*</sup> | 3 (42.9)<sup>*</sup> |
| **MAC capillary deposition** | 230 (89.8) | 84 (96.6)<sup>*</sup> | 23 (57.5)<sup>*</sup> | 25 (86.2) | 82 (98.8)<sup>*</sup> | 10 (100) | 6 (85.7) |
| MAC capillary deposition with perifascicular pattern | 162 (63.3) | 65 (74.7)<sup>*</sup> | 11 (27.5)<sup>*</sup> | 7 (24.1)<sup>*</sup> | 65 (78.3)<sup>*</sup> | 9(90.0) | 5(71.4) |
| Distinct MAC capillary deposition in perifascicular area | 155(60.5) | 62 (71.3)<sup>*</sup> | 8 (20.0)<sup>*</sup> | 9 (31.0)<sup>*</sup> | 63 (75.9)<sup>*</sup> | 8 (80.0) | 5(71.4) |
| **MAC sarcolemmal deposition** | 68(26.6) | 14 (16.1)<sup>*</sup> | 27 (67.5)<sup>*</sup> | 2 (6.9)<sup>*</sup> | 16 (19.3) | 5 (50.0) | 4 (57.1) |
| MAC sarcolemmal deposition with perifascicular pattern | 32 (12.5) | 8 (9.2) | 20 (50.0)<sup>*</sup> | 0 | 2 (2.4)<sup>*</sup> | 2 (20.0) | 0 |

**Abbreviations:** DM, dermatomyositis; anti-TIF1-γ , anti-transcription intermediary factor 1-gamma antibody; anti-Mi-2, anti-complex nucleosome remodeling histone deacetylase antibody; anti-MDA5, anti-melanoma differentiation-associated gene 5; anti-NXP-2, anti-nuclear matrix protein 2; anti-SAE, anti-small ubiquitin-like modifier activating enzyme; PFA, perifascicular atrophy; CD, cluster of differentiation; NA, not available; ALP, alkaline phosphatase; COX, cytochrome C oxidase; ACP, acid phosphatase; MxA, myxovirus resistant protein A; HLA, human leukocyte antigen; MAC, membrane attack complex
Continuous data are shown as mean ± SD; categorical data are shown as number (percentage).
<sup>a</sup>Not evaluated in 1 patient due to insufficient tissue for additional staining
<sup>b</sup>Not evaluated in 1 patient due to tissue artifacts
<sup>c</sup>Not evaluated in 3 patients due to tissue artifacts
<sup>d</sup>Not evaluated in 2 patients due to tissue artifacts
<sup>\*</sup> $p < 0.05$ vs. other antibody subtypes (red- positive association, blue-negative association)
<sup>†</sup> $p < 0.05$ vs. normal specimen (red- positive association, blue-negative association)

### Muscle pathology re-evaluation by histology domains

The muscle fiber domain scores and inflammatory domain scores were significantly different among DMSA subtypes (*P*<.001) (Figure 2A,B). Compared with the group of other DMSAs, muscle biopsies from anti-Mi-2 DM showed significantly higher muscle fiber domain scores (4.9±2.1 vs. non-Mi-2 2.6±2.0, *P*<.001) and higher inflammatory domain scores (8.0±3.0 vs. non-Mi-2 6.3±3.3, *P*=.002) (Table 2). Anti-MDA5 DM showed lower muscle fiber domain score (1.1±1.6 vs. non-MDA5 3.2±2.1, *P*<.001), and lower inflammatory domain score (3.7±2.4 vs. non-MDA5 6.9±3.2, *P*<.001). In normal children, the capillary-to-muscle fiber ratio (CFR) was lower than the ratio in adults.^25^ Thus, we evaluated the vascular domain in juvenile and adult patients separately. The CFR value in all adult DM (0.7±0.3) was lower than the value in 12 normal adult muscle biopsy (1.1±0.1, *P*<.001).^11^ The CFR did not significantly differ among DMSA subtypes (Figure 2C,D). Anti-Mi-2 DM were more commonly associated with increased ALP activity in the perimysium (70.0% vs. non-Mi-2 29.5%, *P*<.001).

**Figure 2.**
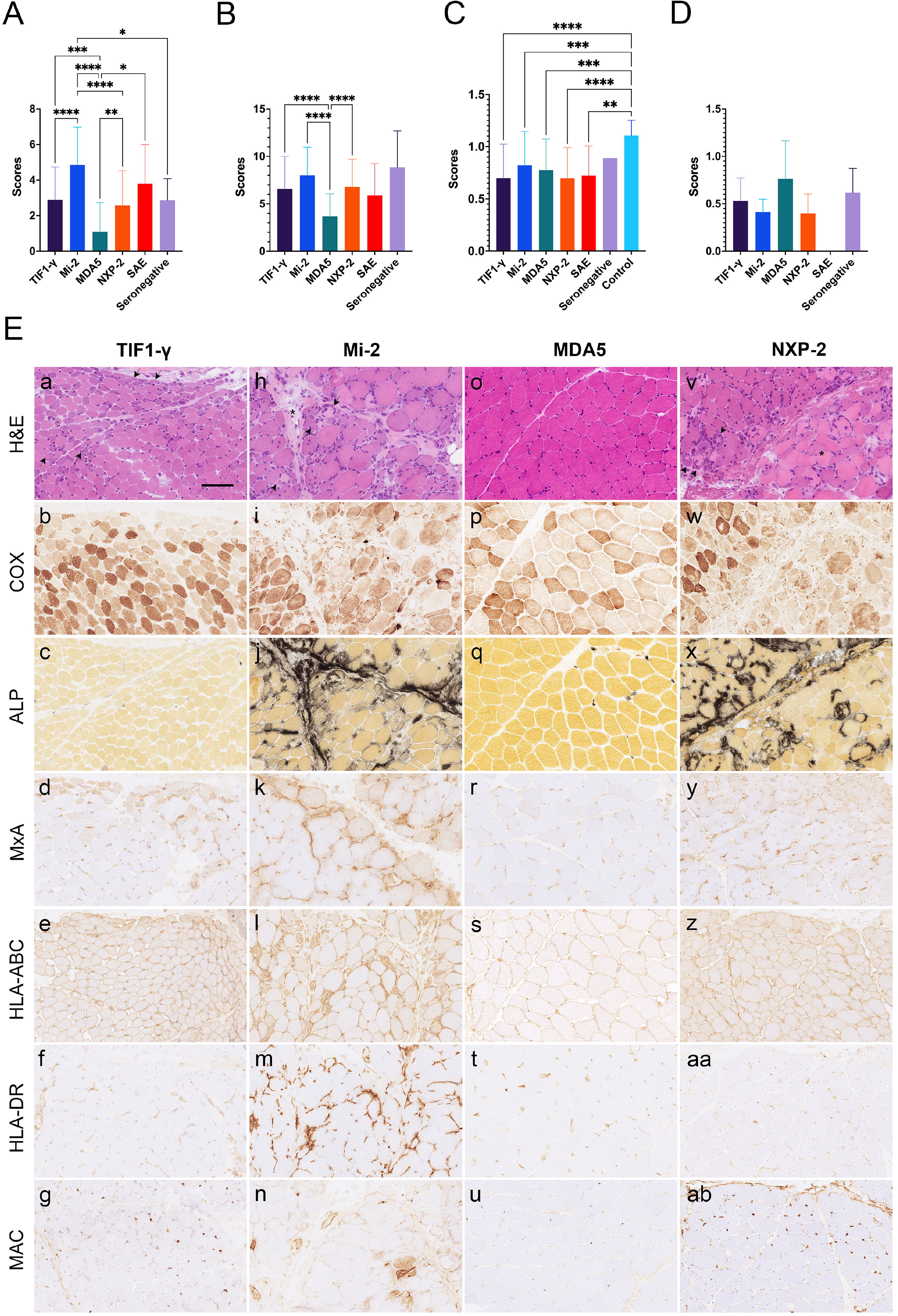
Pathology domains and representative common pathology features among DMSA subtypes. **A. Muscle fiber domain:** anti-Mi-2 DM presented a significantly higher muscle fiber domain score than anti-TIF1-γ, anti-MDA5, anti-NXP-2, and seronegative DM. **B. Inflammatory domain:** anti-MDA5 DM was associated with a lower inflammatory domain score than anti-TIF1-γ, anti-Mi-2, and anti-NXP-2 DM. **C. Vascular domain:** adult capillary-myofiber ratio. The ratio in adult DM was significantly lower than controls. The ratio was not distinctively different among DMSA subtypes. **D. Vascular domain:** juvenile capillary-myofiber ratio. The ratio in juvenile DM was not different across DMSA subtypes. **E. Representative common pathology features among major DMSA subtypes:** anti-TIF1-γ (a-g), anti-Mi-2 (h-n), anti-MDA5 (o-u), and anti-NXP-2 DM (v-ab).On hematoxylin and eosin staining, anti-TIF1-γ DM showed perifascicular atrophy with vacuolated/punched-out fibers (a, black arrow); anti-Mi-2 was associated with perifascicular necrosis (h); anti-MDA5 with near normal appearance (o); and anti-NXP with microinfarction (v, asterisk). central necrotic-peripheral regenerating fibers (CNPR) were present in anti-Mi2 and anti-NXP2 subtypes (black arrows). Decreased COX activity at the periphery of fascicles was prominent in the anti-TIF1-γ DM subtype(b). In anti-MDA5 DM, the COX activity was within normal limits (p). Increased perimysial ALP activity was prevalent in the anti-Mi-2 (j) and anti-NXP-2 DM subtypes (x). MxA staining was present in a perifascicular pattern (d and k), scattered/diffuse (r and y), or mixed pattern (not shown). HLA-ABC was expressed in all MxA-positive DM; perifascicular enhancement was frequent in anti-TIF1-γ DM (e). The pattern was more diffuse in the anti-MDA5-DM subtype. HLA-DR positivity was uncommon (f, m, t, aa – negative). Deposition of MAC on capillaries were prominent in the anti-TIF1-γ (g) and anti-NXP-2 DM (ab) subtypes and was common in the anti-MDA5 DM subtype (u). Anti-Mi-2 DM was frequently associated with sarcolemmal MAC expression (n). **Note:** bar=mean±SD, ANOVA with Dunnette’s T3 multiple comparison ^*^p<0.0332, ^**^p<0.0021, ^***^p<0.0002, ^****^p<0.0001 **Abbreviations:** CK, creatine kinase; DM, dermatomyositis; DMSA, dermatomyositis-specific antibody; COX, cytochrome C oxidase; ALP, alkaline phosphatase; MxA, myxovirus resistant protein A; HLA (human leukocyte antigen); MAC, membrane attack complex

### Histological features of interest

The characteristic features of each DM subtype are shown in Figures 2 and 3. Compared with non-TIF1-γ DM, anti-TIF1-γ DM was significantly associated with vacuolated/punched-out fiber (64.7% vs. non-TIF1-γ 34.3%, *P*<.001). Vacuolated/punched-out fiber was significantly associated with anti-TIF1-γ DM with malignancy (74.3% vs. non-TIF1-γ 15.8%, *P*<.001). The following features were significantly associated with anti-Mi-2 DM: PFA (67.5% vs. non-Mi-2 46.3%, *P*=.02), PFN (52.5% vs. non-Mi-2 3.3%, *P*<.001), CD8 infiltration in non-necrotic fibers (7.5% vs. non-Mi-2 0.9%, *P*=.03), ACP/CD68 infiltration in non-necrotic fiber (20.0% vs. non-Mi-2 2.8%, *P*<.001), CD20 aggregation (28.2% vs. non-Mi-2 13.0%, *P*=.03), and the presence of CNPR (45.0% vs. non-Mi-2 20.8%, *P*=.002). Muscle biopsies from anti-MDA5 DM showed negative associations with PFA (24.1%, vs. non-MDA5 52.9%, *P*=.005), decreased COX activity in perifascicular areas (17.2% vs. non-MDA5 50.0%, *P*<.001), perivascular inflammatory cell infiltration (13.8% vs. non-MDA5 48.0%, *P*<.001), and vacuolated fiber (20.7% vs. non-MDA5, 47.6%, *P*=.009). Anti-NXP2 DM was significantly associated with microinfarction (26.5% vs. non-NXP-2 9.3%, *P*<.001), and this finding was significant for both adult and juvenile patients. While there was a distinctive difference between the percentages of microinfarction affecting adults (12.2%) and juvenile DM patients (22.4%, *P*=.05), the percentage of microinfarctions in adults (20.8%) and juvenile anti-NXP-2 DM (34.3%) was not significantly different (*P*=.21). In our study, anti-SAE DM did not show any distinctive association with the above-mentioned items. Seronegative DM was significantly associated with perivascular inflammation (85.7% vs. seropositive 43.0%, *P*=.05) and vasculitis (57.1% vs. seropositive 14.1%, *P*=.01).

**Figure 3.**
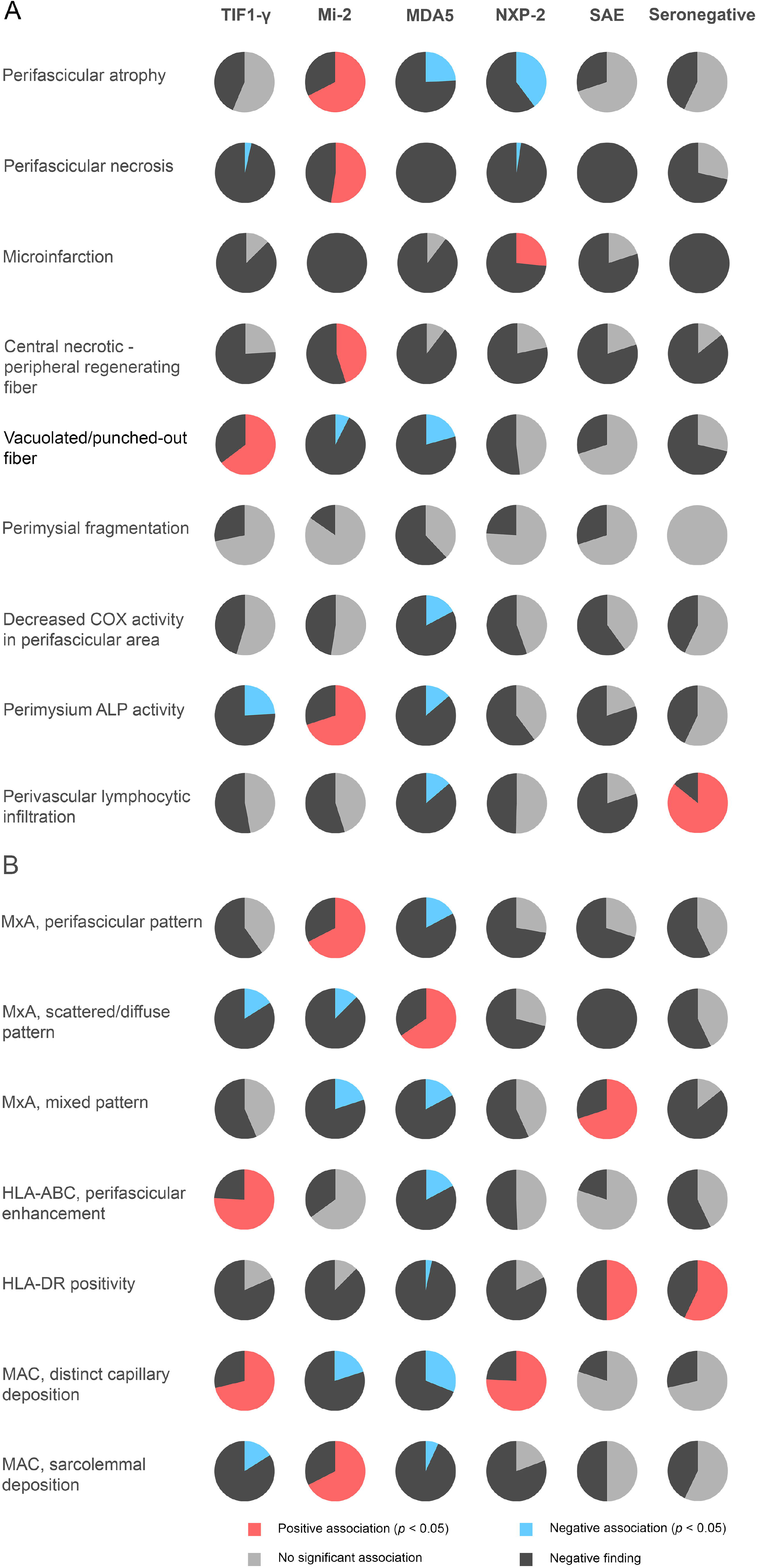
Proportions of histological and immunohistochemical features of interest. **A. Histological features of interest compared with other DMSA subtypes**. **B. Immunohistological features of interest compared with other DMSA subtypes**.

CNPR fibers were commonly present in muscle biopsies with microinfarction (57.9% vs. 18.8% *P*<.001) and 36.8% (14/63) of CNPR were present in the infarcted region (eFigure 2).

### Immunohistochemical features of interest

All muscle biopsies in this study expressed both MxA and HLA-ABC. MxA expression showed 37.5% pure perifascicular staining pattern, 25.4% scattered/diffuse pattern, and 37.1% mixed pattern. Pure perifascicular MxA staining was significantly associated with anti-Mi-2 DM (67.5% vs. non-Mi-2 31.9%, *P*<.001), while a pure scattered/diffuse staining pattern was significantly associated with anti-MDA5 DM (65.5% vs. non-MDA5 20.3%, *P*<.001). HLA-DR positivity, either scattered or perifascicular patterns were significantly associated with anti-SAE (50.0% vs. non-SAE 8.9%, *P*=.002) and seronegative DM (42.9% vs. seropositive 9.6%, *P*=.03). Anti TIF1-γ and anti-NXP-2 DM was significantly associated with capillary MAC deposition (96.6% vs. non-TIF1-γ 86.4%, *P*=.009 and 98.8% vs. non-NXP-2 85.6%, *P*<.001), capillary MAC deposition with perifascicular pattern (74.7% vs. non-TIF1-γ 57.4%, *P*=.007 and 78.3% vs non-NXP-2 56.1%, *P*<.001), and distinct capillary MAC deposition in perifascicular areas (71.3% vs. non-TIF1-γ 55.0%, *P*=.02 and 75.9% vs. non-NXP-2 53.2%, *P*<.001). Distinct capillary MAC deposition in anti-TIF1-γ DM was not significantly associated with malignancy (69.4% vs. non-malignancy associated TIF-1-y 72.6%, *P*=0.81). Sarcolemmal MAC deposition of any pattern or with some area showing perifascicular pattern was significantly present in anti-Mi-2 DM (67.5% vs. non-Mi-2 19.0%, *P*<.001 and 50.0% vs. non-Mi-2 5.6%, *P*<.001, respectively).

Because the features in anti-TIF1-γ and anti-NXP-2 DM tended to be overlapping, we compared the features in both subtypes (eTable 4). Anti-TIF1-γ DM was associated with perifascicular atrophy (*P*=.03), vacuolated/punched out fiber (*P*=.04), and HLA-ABC expression with perifascicular enhancement (*P*=.05). Anti-NXP-2 DM was associated with ALP activity in PFA (*P*=.03) and microinfarction (*P*=.03).

## Discussion

This was an extensive study on serological-pathological correlations of different DMSA subtypes. The sarcoplasmic MxA expression was used as a diagnostic finding of DM on muscle pathology as it has proven to be highly sensitive (71-77%) and specific (98-100%) for DM.^5, 6^ We also confirmed that the presence of MxA expression was more sensitive than PFA for the diagnosis of DM (100% vs. 49.6%).^5^ However, as only MxA with perifascicular pattern is mentioned in the 2018 ENMC-DM classification, this may result in underdiagnosing or misclassification of 25.4% of DM muscle biopsies. We thus propose that sarcoplasmic MxA expression in non-necrotic/regenerating fibers *per se* should be regarded as diagnostic of DM, regardless of the pattern of the distribution of MxA-positive fibers. Using MxA expression as a criterion, we identified 7 seronegative DM that warrant further study and have yet-to-be-identified as DMSAs or as truly seronegative.

We regarded DMSD as a form of DM.^16^ Thus, unlike the 2018 ENMC-DM classification, which requires identification of cutaneous DM features either clinically or pathologically,^2^ we included all qualified muscle biopsies in our serological-pathological criteria, regardless of the cutaneous features. In this extended cohort,^16^ nineteen anti-NXP-2 antibody-positive patients (19/83, 22.9%) presented no skin lesion of any kind at the time of diagnosis and were classified as DMSD. The proportion was comparable with cases recently reported in a separate study (8/76, 10.5%, *P*=.06).^22^

In addition to the DMSA-associated clinical phenotypes described above, in our cohort, anti-TIF1-γ and anti-Mi-2 were significantly associated with adult patients while anti-NXP-2 was significantly associated with juvenile patients. Anti-SAE DM was associated with elderly patients and a low CK level and showed a trend for DM skin lesions, muscle weakness, dysphagia, and concurrent malignancy, albeit not significantly, which may be at least partly due to the limited number of patients in this study. Seronegative DM was associated with juvenile patients with less frequent DM skin lesions. However, clinical information was limited owing to the retrospective nature of the study.

We identified significantly frequent pathological features for each DM subtype classified by DMSAs; some were previously described in smaller studies. Histological features in anti-Mi-2 DM were compatible with those previously described in immune myopathy with perimysial pathology (IMPP).^11,18^ By comparing each DMSA with other subtypes, anti-Mi-2 was significantly associated with PFA,^11,13^ PFN,^11,18^ PM-ALP,^11,18^ inflammatory features,^11-13^ perifascicular MxA expression, and sarcolemmal MAC expression.^11,13,18^ We confirmed that anti-TIF1-γ DM was significantly associated with vacuolated fibers,^8^ HLA-ABC expression with perifascicular enhancement and distinctive capillary MAC deposition.^8^ Further, vacuolated/punched-out fibers were significantly associated with malignancy-associated anti-TIF1-γ DM.^8^ However, we could not demonstrate a significant association between distinctive capillary MAC deposition and malignancy. Although anti-TIF1-γ and anti-NXP2 DM were very similar, anti-TIF-γ DM were more significantly associated with PFA and HLA-ABC expression with perifascicular enhancement, features described for DM with vasculopathy (DM-VP).^26, 27^

Anti-NXP-2 was distinctively associated with microinfarctions, features described in regional ischemic immune myopathy (RIIM).^26^ Although few cases have been reported describing muscle ischemia or clustered necrosis in juvenile anti-NXP-2 DM, ^18, 19^ we provide the first evidence that adult and juvenile anti-NXP-2 DM were equally affected by microinfarction. Since the percentage of microinfarctions in adult and juvenile anti-NXP-2 DM were not significantly different, this phenomenon could not simply be explained by different CFRs between the two age groups. The percentage of MAC deposition on capillaries in anti-NXP-2 DM was virtually equal to the percentage in anti-TIF1-γ DM but the percentages of microinfarction were significantly different. Thus, microinfarction could not be explained by complement deposition and warrants further study.

CNPR fibers are peculiar necrotic-regenerating fibers, described as rare in muscular dystrophies,^28^ but their presence has never been fully studied. In our experience, CNPR was not observed in 140 muscle biopsies with genetically confirmed muscular dystrophies (eTable 5), albeit these muscular dystrophies may sometimes demonstrate myositis-mimicking pathology.^29^ It appears that crescent-shaped small regenerating fibers are present at the periphery of necrotic fibers although detailed morphological analysis could not be performed as samples were not available for electron microscopy for the patients with CNPR fibers.

In muscular dystrophies and necrotizing myopathies, necrotic fibers are often invaded by macrophages which are believed to clean up the liquefied sarcoplasm while regeneration is initiated in parallel. In our study, CNPR fibers were significantly associated with microinfarction and some of the neighboring fibers contained lipid droplets or cytoplasmic bodies. These findings, together with their hypoxia-inducible factor expression suggested that they reflect hypoxic injury. Of note, infarcted fibers do not usually accompany macrophage invasion, which may partly explain why CNPR fibers did not undergo phagocytosis. Interestingly, CNPR fibers were more commonly present in anti-Mi-2 DM which was not associated with microinfarction. Further studies are necessary to better understand whether CNPR fibers in anti-Mi-2 are developed through a mechanism different from those in anti-NXP-2 DM. Anti-MDA5 DM was associated with near normal pathology^18,19,30^ and scattered/diffuse MxA positivity ^6^, while anti-SAE DM were distinctively associated with HLA-DR expression. Seronegative DM showed a trend toward inflammation and was associated with significant HLA-DR expression. Since HLA-DR expression is a marker of the type II interferon (IFN2)^31^, there may well be more prominent IFN2 activation in these antibody subtypes.

In conclusion, our study demonstrated distinctive myopathological features associated with DMSA subtypes, which may well indicate the presence of different underlying pathobiological mechanisms. Inclusion of these features into the classification criteria would not only increase diagnostic yield but also help classify DM subtypes.

## Supporting information

eTable1

eTable2

eTable3

eTable4

eTable5

eFigure1

eFigure2

## Data Availability

Any anonymized data relevant to the study not published within the article will be shared at the request of any qualified investigator.

## Acknowledgements

The authors thank Kazu Iwazawa, Kaoru Tatezawa, Naho Fushimi, Miyuki Matsuda, and Hisayoshi Nakamura for their technical assistance.

## Access to Data Statement

JT and IN had full access to all the data in the study and takes responsibility for the integrity of the data and the accuracy of the data analysis.

