## supplement for "Dermatomyositis: Muscle Pathology According to Antibody Subtypes"

**eMethods**

**Histochemical and immunohistochemical staining**

Histochemical and immunohistochemical staining used in this study, included hematoxylin and eosin (H&E), modified Gomori trichrome (mGT), acid phosphatase (ACP), alkaline phosphatase (ALP), cytochrome C oxidase (COX), Oil Red O, nicotinamide adenine dinucleotide tetrazolium reductase (NADH-TR), MxA (Mx 1/2/3, Santa Cruz Biotechnology), class I human major histocompatibility complex (MHC) (HLA-ABC; clone W6/32, Thermo Fisher Scientific), MHC class II (HLA-DR; clone B308, Affinity BioReagents), membrane attack complex (MAC; C5b-9, clone aE11, Dako), neonatal myosin heavy chain (MHCn; clone WB-MHCn, Leica), utrophin (clone DRP3/20C65, Novocastra), CD3 (polyclonal, Abcam), CD8 (clone DK25, Dako), CD20 (clone L26, eBioscience), and CD68 (clone KP1, Dako).

**Histochemical and immunohistochemical evaluation**

Histochemical and immunohistochemical staining was performed for routine diagnostic purposes.

JT performed pathology evaluation and was blinded to the antibody result at the time of evaluation. Histochemical and immunohistochemical stained slides, prepared at the time of pathological diagnosis at the NCNP were evaluated based on 4 domains (muscle fiber, inflammatory, vascular, and connective tissue domains) using the same scoring system as our previous studies modified from the pathology scoring system originally used for juvenile DM.^10,14^ The scoring system for the muscle fiber domain was based on the presence and extent of necrotic fibers, regenerating fibers, atrophic fibers separated from the perifascicular area, and fibers with internalized nuclei; the inflammatory domain consists of CD3-, CD20-, and CD68-positive inflammatory cells in the endomysium and perimysium, and the presence of perivascular inflammatory cell infiltration; the vascular domain consisted of the capillary to muscle fiber ratio; and the connective tissue domain consisted of endomysium fibrosis and perimysium pathology which included fragmentation and alkaline phosphatase expression.^10^ Sarcoplasmic MAC deposition and MHCn expression were used as supporting findings to identify and confirm necrotic and regenerating fibers, respectively. Utrophin was used as a surrogate endothelial marker to evaluate the capillary-to-muscle fiber ratio.^10^
