## Supplementary material for "Dermatomyositis: Muscle Pathology According to Antibody Subtypes": eTable1

**eTable 1**

**Pathology domains and scoring/evaluation criteria used for muscle biopsy assessment**

| **Pathology domain** | **Scoring/evaluation criteria** |
| --- | --- |
| **Muscle fiber domain** | Maximum score = 8 |
| Necrotic fiber | 0 = absence, 1 = isolated/sporadic 1–3 per 20x, 2 = scattered ≥4 or more in adjacent fibers, or clustered, or >3 per 20x |
| Regenerating fiber | 0 = <6 per 20x, 1 = ≥6 per 20x |
| Atrophic fibers away from perifascicular area | 0 = absence, 1 = isolated/sporadic 1–3 per 20x, 2 = scattered ≥ 4 or more in adjacent fibers, or clustered, or >3 per 20x |
| Perifascicular atrophy | 0 = absence, 1 = affecting 1-2 fascicle(s), 2 = affecting ≥ 2 fascicles |
| Fiber with internalized nuclei >3% | 0 = ≤3%, 1 = >3% |
| **Inflammatory domain** | Maximum score = 13 |
| Endomysial and perimysial CD3+ cell infiltration | 0 = none or <4 cells in 20x field, 1 = 4–20 cells in a 20x field or a cluster of ≥10 cells, 2 = ≥2 clusters in whole biopsy and/or diffusely infiltrating i.e. >20 cells in a 20x field (separate scores for endomysial and perimysial infiltration) |
| Endomysial and perimysial CD20+ cell infiltration |  |
| Endomysial and perimysial CD68+ cell infiltration |  |
| Perivascular inflammatory cell infiltration | 0 = absence, 1 = presence |
| **Vascular domain** | |
| Capillary:fiber ratio | Utrophin-positive capillaries per the number of muscle fibers in 10 randomly photographed areas at 20x or at least 5 areas at 20x in the case of small biopsy |
| **Connective tissue domain** | |
| Perimysial connective tissue fragmentation | N = absence, Y = presence |
| Increased alkaline phosphatase activity in perimysium | N = absence, Y = presence |
| Endomysial fibrosis | N = absence, Y = presence |

Abbreviation: N = no; Y= yes
