## Supplementary material for "Dermatomyositis: Muscle Pathology According to Antibody Subtypes": eTable2

**eTable 2**

**Disease duration and exposure to systemic immunotherapy before biopsy**

|  | All DM | Anti-TIF1-γ | Anti-Mi-2 | Anti-MDA5 | Anti-NXP-2 | Anti-SAE | Seronegative |
| --- | --- | --- | --- | --- | --- | --- | --- |
|  | (n = 256) | (n = 87) | (n = 40) | (n = 29) | (n = 83) | (n = 10) | (n = 7) |
| Disease duration (months) | 6.7± 13.4 | 8.1 ± 13.4 | 5.0 ± 4.0 | 4.1± 6.0 | 7.2 ± 18.4 | 4.7 ± 3.2 | 7.3 ± 5.6 |
| **Systemic immunotherapy** |  |  |  |  |  |  |  |
| Within 6 weeks before biopsy^a^ | 48 (18.8) | 11 (12.6) | 9 (22.5) | 9 (31.0) | 15 (18.1) | 3 (30.0) | 1(14.3) |
| **Biopsy site** |  |  |  |  |  |  |  |
| ***Proximal muscle upper extremity*** | | | | | | | |
| Biceps brachii | 132 (51.6) | 37 (42.5)* | 26 (65.0) | 12 (41.4) | 45 (54.2) | 7 (70.0) | 5 (71.4) |
| Triceps brachii | 8 (3.1) | 5 (5.7) | 2 (5.0) | 0 | 1 (1.2) | 0 | 0 |
| Deltoid | 23 (9.0) | 10 (11.5) | 2 (5.0) | 4 (13.8) | 7 (8.4) | 0 | 0 |
| ***Proximal muscle, lower extremity*** | | | | | | | |
| Quadriceps, nos | 23 (9.0) | 10 (11.5) | 4 (10.0) | 4 (13.8) | 4 (4.8) | 1 (10.0) | 0 |
| Rectus femoris | 26 (10.2) | 13 (14.9) | 2 (5.0) | 0 | 10 (12.0) | 1 (10.0) | 0 |
| Vastus intermedius | 1 (0.4) | 1 (1.1) | 0 | 0 | 0 | 0 | 0 |
| Vastus lateralis | 33 (12.9) | 9 (10.3) | 3 (7.5) | 7 (24.1) | 13 (15.7) | 1 (10.0) | 0 |
| Vastus medialis | 1 (0.4) | 0 | 1 (2.5) | 0 | 0 | 0 | 0 |
| Sartorius | 1 (0.4) | 0 | 0 | 1 (3.4) | 0 | 0 | 0 |
| Semitendinosus | 2 (0.8) | 1 (1.1) | 0 | 0 | 1 (1.2) | 0 | 0 |
| Biceps femoris | 1 (0.4) | 0 | 0 | 0 | 0 | 0 | 1 (14.3) |
| ***Distal muscle, lower extremity*** | | | | | | | |
| Gastrocnemius | 1 (0.4) | 0 | 0 | 0 | 0 | 0 | 1 (14.3) |
| Tibialis anterior | 4 (1.6) | 1 (1.1) | 0 | 1 (3.4) | 2 (2.4) | 0 | 0 |

**Abbreviations:** DM, dermatomyositis; anti-TIF1-γ, anti-transcription intermediary factor 1-gamma antibody; anti-Mi-2, anti-complex nucleosome remodeling histone deacetylase antibody; anti-MDA5, anti-melanoma differentiation-associated gene 5; anti-NXP-2, anti-nuclear matrix protein 2; anti-SAE, anti-small ubiquitin-like modifier activating enzyme

Continuous data are shown as mean ± SD; categorial data are shown as number (percentage).

**p* < 0.05 (compare with other antibody subtypes
