## Supplementary material for "Dermatomyositis: Muscle Pathology According to Antibody Subtypes": eTable3

**Agglomerative hierarchical clustering (AHC): Member and variable of classes**

| **Variables** | **Class 1** | | **Class 2** | | **Class 3** | | **Class 4** | | **Class 5** | | **Class 6** | |
| --- | --- | --- | --- | --- | --- | --- | --- | --- | --- | --- | --- | --- |
|  | (n = 32) | | (n = 37) | | (n = 52) | | (n = 67) | | (n = 37) | | (n = 28) | |
|  | n | % | n | % | n | % | n | % | n | % | n | % m |
| Anti-TIF1-γ (total = 85)^a^ | 2 | 6.3 (2.4) | 12 | 32.4 (14.1) | 7 | 13.5 (8.2) | 31 | 46.3 (36.5) | 25 | 67.6 (29.4) | 8 | 28.6 (9.4) |
| Anti-Mi-2 (total = 39)^b^ | 29 | 90.6 (74.4) | 2 | 5.4 (5.1) | 1 | 1.9 (2.6) | 0 | 0 | 7 | 18.9 (17.9) | 0 | 0 |
| Anti-MDA5 (total = 29) | 0 | 0 | 0 | 0 | 21 | 40.4 (72.4) | 4 | 6.0 (13.8) | 4 | 10.8 (13.8) | 0 | 0 |
| Anti-NXP-2(total = 83) | 1 | 3.1 (1.2) | 23 | 62.2 (27.7) | 23 | 44.2 (27.7) | 32 | 47.8 (38.6) | 0 | 0 | 4 | 14.3 (4.8) |
| Anti-SAE(total = 10) | 0 | 0 | 0 | 0 | 0 | 0 | 0 | 0 | 1 | 2.7 (10.0) | 9 | 32.1 (90.0) |
| Seronegative (total =7) | 0 | 0 | 0 | 0 | 0 | 0 | 0 | 0 | 0 | 0 | 7 | 25.0(100) |
| Perimysium ALP activity, increased (total = 91) | 28 | 87.5 (30.8) | 19 | 51.4 (20.9) | 1 | 1.9 (1.1) | 27 | 40.3 (29.7) | 3 | 8.1 (3.3) | 13 | 46.4 (14.3) |
| PFA (total = 125) | 24 | 75.0 (19.2) | 34 | 91.9 (27.2) | 6 | 11.5 (4.8) | 20 | 29.9 (16.0) | 18 | 48.6 (14.4) | 23 | 82.1 (18.4) |
| PFN (total = 28) | 22 | 68.8 (78.6) | 0 | 0 | 0 | 0 | 2 | 3.0 (7.1) | 2 | 5.4 (7.1) | 2 | 7.1 (7.1) |
| Vacuolated/punched-out fibers (total = 113) | 2 | 6.3 (2.0) | 26 | 70.3 (26.5) | 6 | 11.5 (6.1) | 34 | 50.7 (34.7) | 14 | 37.8 (14.3) | 16 | 57.1 (16.3) |
| Decreased COX activity in perifascicular area (total = 116) | 17 | 53.1 (14.7) | 34 | 91.9 (29.3) | 8 | 15.4 (6.9) | 25 | 37.3 (21.6) | 14 | 37.8(12.1) | 18 | 64.3 (15.5) |
| Perivascular inflammatory cell infiltration (total = 111) | 16 | 50.0 (14.4) | 32 | 86.5 (28.8) | 7 | 13.5 (6.3) | 26 | 38.8 (23.4) | 13 | 35.1 (11.7) | 17 | 60.7 (15.3) |
| Vasculitis (total = 39) | 9 | 28.1 (23.1) | 16 | 43.2 (41.0) | 2 | 3.8 (5.1) | 3 | 4.5 (7.7) | 2 | 5.4 (5.1) | 7 | 25.0 (17.9) |
| CD8 infiltration in non-necrotic fiber (total = 5) | 3 | 9.4 (60.0) | 0 | 0 | 0 | 0 | 0 | 0 | 0 | 0 | 2 | 7.1 (40.0) |
| ACP/CD68 infiltration in non-necrotic fiber (total = 14) | 9 | 28.1 (64.3) | 1 | 2.7 (7.1) | 0 | 0 | 2 | 3.0 (14.3) | 0 | 0 | 2 | 7.1 (14.3) |
| CD20 aggregation (total =39) | 10 | 31.3 (25.6) | 15 | 40.5 (38.5) | 0 | 0 | 4 | 6.0 (10.3) | 3 | 8.1 (7.7) | 7 | 25.0 (17.9) |
| Microinfarction(total = 38) | 0 | 0 | 4 | 10.8 (10.5) | 2 | 3.8 (5.3) | 30 | 44.8 (78.9) | 0 | 0 | 2 | 7.1 (5.3) |
| CNPR(total = 62) | 19 | 59.4 (30.6) | 3 | 8.1 (4.8) | 2 | 3.8 (3.2) | 31 | 46.3 (50.0) | 2 | 5.4 (3.2) | 5 | 17.9 (8.1) |
| MxA, perifascicular pattern (total = 94) | 20 | 62.5 (21.3) | 13 | 35.1 (13.8) | 12 | 23.1 (12.8) | 2 | 3.0 (2.1) | 34 | 91.9 (36.2) | 13 | 46.4 (13.8) |
| MxA, scattered/ diffuse pattern (total = 65) | 3 | 9.4 (4.6) | 2 | 5.4 (3.1) | 35 | 67.3 (53.8) | 19 | 28.4 (29.2) | 3 | 8.1 (4.6) | 3 | 10.7 (4.6) |
| MxA, mixed pattern(total = 95) | 9 | 28.1 (9.5) | 22 | 59.5 (23.2) | 5 | 9.6 (5.3) | 47 | 70.1 (49.5) | 0 | 0 | 12 | 42.9 (12.6) |
| HLA-ABC, diffuse pattern (total =105) | 12 | 37.5 (11.4) | 1 | 2.7 (1.0) | 48 | 92.3 (45.7) | 38 | 56.7 (36.2) | 0 | 0 | 6 | 21.4 (5.7) |
| HLA-ABC, perifascicular enhancement (total = 148) | 20 | 62.5 (13.5) | 36 | 97.3 (24.3) | 4 | 7.7 (2.7) | 29 | 43.3 (19.6) | 37 | 100(25.0) | 22 | 78.6 (14.9) |
| HLA-DR positivity (total = 46) | 5 | 15.6 (10.9) | 5 | 13.5 (10.9) | 1 | 1.9 (2.2) | 12 | 17.9 (26.1) | 2 | 5.4 (4.3) | 21 | 75.0 (45.7) |
| HLA-DR, perifascicular pattern (total = 27) | 1 | 3.1 (3.7) | 0 | 0 | 0 | 0 | 7 | 10.4 (25.9) | 0 | 0 | 19 | 67.9 (70.4) |
| MAC, sarcolemmal deposition (total = 67) | 25 | 78.1 (37.3) | 15 | 40.5 (22.4) | 4 | 7.7 (6.0) | 6 | 9.0 (9.0) | 2 | 5.4 (3.0) | 15 | 53.6 (22.4) |
| MAC, sarcolemmal deposition, perifascicular pattern (total = 31) | 21 | 65.6 (67.7) | 3 | 8.1 (9.7) | 1 | 1.9 (3.2) | 2 | 3.0 (6.5) | 0 | 0 | 4 | 14.3 (12.9) |
| MAC, capillary deposition, perifascicular pattern (total = 161) | 10 | 31.3 (6.2) | 33 | 89.2 (20.5) | 14 | 26.9 (8.7) | 60 | 89.6 (37.3) | 18 | 48.6 (11.2) | 26 | 92.9 (16.1) |
| Distinct MAC capillary deposition, perifascicular area (total = 154) | 9 | 28.1 (5.8) | 31 | 83.8 (20.1) | 15 | 28.8 (9.7) | 62 | 92.5 (40.3) | 14 | 37.8 (9.1) | 23 | 82.1 (14.9) |

**Abbreviations:** DM, dermatomyositis; anti-TIF1-γ, anti-transcription intermediary factor 1-gamma antibody; anti-Mi-2, anti-complex nucleosome remodeling histone deacetylase antibody; anti-MDA5, anti-melanoma differentiation-associated gene 5; anti-NXP-2, anti-nuclear matrix protein 2; anti-SAE, anti-small ubiquitin-like modifier activating enzyme; CD, cluster of differentiation; ALP, alkaline phosphatase; PFA, perifascicular atrophy; PFN, perifascicular necrosis; COX, cytochrome C oxidase; ACP, acid phosphatase; CNPR, central necrotic-peripheral regenerating fiber; MxA, myxovirus resistant protein A; HLA, human leukocyte antigen; MAC, membrane attack complex
