## Supplementary material for "Dermatomyositis: Muscle Pathology According to Antibody Subtypes": eTable4

**eTable 4 Histological and immunohistochemical features in anti-TIF1-γ DM vs. Anti-NXP2 DM**

|  | **Anti-TIF1-γ DM** | **Anti-NXP-2 DM** | ***p* value** |
| --- | --- | --- | --- |
|  | (n = 87) | (n = 83) |  |
| Perimysial fragmentation† | 61(71.8)^a^ | 63(75.9) | 6.0e-1 |
| Perimysial ALP activity, increased† | 21(24.1) | 33(39.8)* | 3.3e-2 |
| Endomysial fibrosis | 12(14.1)^a^ | 10(12.0) | 3.2e-1 |
| Perifascicular atrophy‡ | 49(56.3)* | 33(39.8) | 3.3e-2 |
| Perifascicular necrosis† | 3(3.4) | 2(2.4) | 1.0 |
| Decreased COX activity in perifascicular area‡ | 47(54.7)^b^ | 37(44.6) | 2.2e-1 |
| Perivascular inflammatory cell infiltration | 41(47.1) | 42(50.6) | 8.8e-1 |
| Vasculitis | 10(11.5) | 15(18.1) | 2.8e-1 |
| CD8 infiltration in non-necrotic fiber | 2(2.3) | 0 | 5.0e-1 |
| ACP/CD68 infiltration in non-necrotic fiber | 4(4.6) | 1(1.2) | 3.7e-1 |
| CD20 aggregation‡ | 10(11.5) | 13(15.7) | 5.0e-1 |
| Microinfarction§ | 11(12.6) | 22(26.5)* | 3.2e-2 |
| Microinfarction, adult patients | 10(13.9) | 10(20.8) | 3.3e-1 |
| Microinfarction, juvenile patient | 1(6.7) | 12(34.3) | 7.6e-2 |
| Central necrotic -peripheral regenerating fibers (CNPR) | 21(24.1) | 18(21.7) | 7.2e-1 |
| Vacuolated, “punched out” fiber | 55(64.7)^a^* | 40(48.2) | 4.3e-2 |
| MXA positivity | 87(100.0) | 83(100.0) | 1.0 |
| MXA, perifascicular pattern | 35(40.2) | 23(27.7) | 1.1e-1 |
| MXA, scattered/diffuse | 14(16.1) | 24(28.9) | 6.5e-2 |
| MXA, mixed pattern | 38(43.7) | 36(43.4) | 1.0 |
| HLA-ABC positivity | 87(100.0) | 83(100.0) | 1.0 |
| HLA-ABC, perifascicular enhancement‡ | 66(75.9)* | 41(49.4) | 4.5e-4 |
| HLA-DR positivity | 16(18.4) | 15(18.1) | 1.0 |
| HLA-DR, perifascicular pattern | 8(9.2) | 10(12.0) | 6.2e-1 |
| MAC capillary deposition‡ | 84(96.6) | 82(98.8) | 6.2e-1 |
| MAC capillary deposition with perifascicular pattern‡ | 65(74.7) | 65(78.3) | 5.9e-1 |
| Distinct MAC capillary deposition in perifascicular area‡ | 62(71.3) | 63(75.9) | 6.0e-1 |
| MAC sarcolemmal deposition | 14(16.1) | 16(19.3) | 6.9e-1 |
| MAC sarcolemmal deposition with perifascicular pattern | 8(9.2) | 2(2.4) | 1.0e-1 |

**Abbreviations:** DM, dermatomyositis; anti-TIF1-γ, anti-transcription intermediary factor 1-gamma antibody; anti-Mi-2, anti-complex nucleosome remodeling histone deacetylase antibody; anti-MDA5, anti-melanoma differentiation-associated gene 5; anti-NXP-2, anti-nuclear matrix protein 2; anti-SAE, anti-small ubiquitin-like modifier activating enzyme; PFA, perifascicular atrophy; CD, cluster of differentiation; NA, not available; ALP, alkaline phosphatase; COX, cytochrome C oxidase; ACP, acid phosphatase; MxA, myxovirus resistant protein A; HLA, human leukocyte antigen; MAC, membrane attack complex. Categorial data are shown as number (percentage). ^a^Not evaluate in 2 patients due to tissue artifacts ^b^Not evaluate in 1 patient due to tissue artifacts**p* < 0.05 compare with other antibody subtypes (red- positive association)†Features described in immune myopathies with perimysial pathology (IMPP)‡Features described in dermatomyositis with vascular pathology (DM-VP)§Features described in regional ischemic immune myopathy (RIIM)
