## Supplementary material for "Dermatomyositis: Muscle Pathology According to Antibody Subtypes": eTable5

e**Table 5 CNPR fiber in muscular dystrophies* that could be associated with myositis mimic pathology^29^**

| **Muscular dystrophies** | **n** | **CNPR** |
| --- | --- | --- |
| Anoctaminopathy | 3 | 0 |
| Dysferlinopathy | 50 | 0 |
| FKRP-related myopathy | 9 | 0 |
| FSHD | 47 | 0 |
| Laminopathy | 16 | 0 |
| Sarcoglycanopathy | 15 | 0 |
| Total | 140 | 0 |

Abbreviation: CNPR, central necrotic-peripheral regenerating; FKRP, fukutin-related protein; FSHD, facioscapulohumeral*Immunohistochemically/genetically confirmed from January 2009-September 2019
