## Supplementary material for "Dermatomyositis: Muscle Pathology According to Antibody Subtypes": eFigure1

**A.**

Observations (axes F1 and F2: 29.54 %)

F2 (13.14 %)

| F1 (16.40 %) | MDA5 (Obs) |
| --- | --- |
| -0.776923 | 0.263554 |
| -0.659614 | 0.265552 |
| -0.776923 | 0.263554 |
| -0.0399664 | 0.00206994 |
| -0.419606 | -0.170381 |
| -0.284131 | 0.373045 |
| -0.536384 | 0.0510696 |
| -0.0541311 | -0.251952 |
| -0.776923 | 0.263554 |
| -0.776923 | 0.263554 |
| -0.776923 | 0.263554 |
| -0.776923 | 0.263554 |
| -0.776923 | 0.263554 |
| -0.636617 | 0.0963496 |
| -0.641566 | 0.0817877 |
| -0.489432 | -0.146037 |
| -0.434574 | -0.0357845 |
| -0.776923 | 0.263554 |
| -0.567569 | 0.188324 |
| -0.265763 | 0.278437 |
| 0.048263 | -0.200933 |
| -0.624698 | 0.145549 |
| -0.468042 | 0.153558 |
| -0.677981 | 0.360161 |
| -0.776923 | 0.263554 |
| -0.713104 | 0.130208 |
| -0.624698 | 0.145549 |
| -0.776923 | 0.263554 |
| -0.18349 | 0.0726825 |

| F1 (16.40 %) | Mi2 (Obs) |
| --- | --- |
| 0.306888 | 0.347464 |
| 0.433287 | 0.314678 |
| 0.499044 | 1.13173 |
| -0.308872 | 0.598366 |
| -0.0289415 | 0.557629 |
| 0.478591 | 0.399756 |
| 0.625589 | 0.869732 |
| 0.849558 | 0.871612 |
| 0.29206 | 0.898668 |
| 0.216742 | 0.560076 |
| 0.247073 | 0.702971 |
| 0.119946 | 0.384865 |
| 0.54698 | 0.74589 |
| -0.187436 | 0.429017 |
| -0.128468 | 0.592395 |
| 0.758328 | 1.23279 |
| 0.640923 | 0.715855 |
| -0.0708485 | 0.616662 |
| 0.574504 | 0.700944 |
| 0.921842 | 1.03966 |
| 0.0191113 | 0.155573 |
| 0.00629447 | 0.0531962 |
| 0.511417 | 0.105461 |
| 0.0101296 | 0.495084 |
| 0.474207 | 1.09149 |
| 0.133204 | 0.758624 |
| -0.34423 | 0.475281 |
| 0.0034689 | 0.427163 |
| 0.290796 | 0.891462 |
| 0.409051 | 1.08378 |
| 0.27571 | 0.723454 |
| -0.34423 | 0.475281 |
| -0.123616 | 0.295671 |
| 0.745993 | 0.595764 |
| -0.301215 | 0.381392 |
| 0.53444 | 1.06279 |
| 0.389481 | 0.268857 |
| -0.187436 | 0.429017 |
| 0.790843 | 1.06956 |

| F1 (16.40 %) | NXP2 (Obs) |
| --- | --- |
| 0.530296 | -0.211747 |
| 0.213372 | -0.141922 |
| -0.164857 | -0.0975064 |
| -0.0220519 | -0.489133 |
| -0.181207 | -0.27272 |
| 0.405747 | -0.456454 |
| -0.494733 | 0.212602 |
| -0.496257 | -0.218645 |
| 0.196703 | -0.444283 |
| -0.600857 | 0.0471394 |
| 0.288472 | 0.134353 |
| -0.663308 | 0.155394 |
| -0.244505 | -0.371417 |
| 0.0761302 | -0.290317 |
| -0.0685804 | -0.250526 |
| -0.558708 | -0.11039 |
| -0.494733 | 0.212602 |
| 0.0724326 | -0.428244 |
| 0.501939 | -0.250182 |
| -0.307859 | -0.258936 |
| -0.226526 | -0.352231 |
| -0.558708 | -0.11039 |
| 0.402161 | -0.387252 |
| 0.0736494 | -0.10609 |
| -0.118756 | -0.380975 |
| 0.537455 | -0.242879 |
| -0.365507 | -0.190259 |
| -0.214077 | 0.0464884 |
| -0.0326861 | -0.374057 |
| 0.172272 | -0.136631 |
| 0.237618 | 0.0252241 |
| -0.262415 | -0.152939 |
| -0.622527 | 0.0229554 |
| -0.663308 | 0.155394 |
| -0.494733 | 0.212602 |
| -0.494733 | 0.212602 |
| 0.305334 | -0.240383 |
| -0.599488 | 0.022048 |
| -0.133172 | -0.134174 |
| -0.0513044 | -0.451285 |
| 0.0391409 | -0.0305708 |
| 0.609735 | -0.0867449 |
| 0.616255 | -0.175919 |
| -0.367803 | -0.112244 |
| 0.7003 | -0.199816 |
| 0.101211 | -0.451294 |
| -0.122809 | -0.209251 |
| -0.444788 | -0.0567688 |
| -0.622527 | 0.0229554 |
| -0.274941 | -0.30333 |
| -0.0870246 | -0.143172 |
| -0.46413 | -0.159719 |
| -0.164857 | -0.0975064 |
| -0.0816805 | -0.307487 |
| -0.367803 | -0.112244 |
| -0.223591 | -0.372203 |
| 0.617322 | -0.122824 |
| -0.295555 | -0.10251 |
| 0.171675 | -0.417595 |
| 0.281166 | -0.184858 |
| 0.134572 | -0.208882 |
| 0.428167 | -0.137433 |
| 0.538477 | 0.246478 |
| 0.400052 | -0.132616 |
| 0.00224744 | -0.126107 |
| 0.292116 | -0.453147 |
| -0.0311105 | -0.0491915 |
| -0.221987 | -0.140282 |
| 0.2758 | -0.482742 |
| 0.276877 | -0.399027 |
| 0.562325 | -0.381055 |
| 0.137383 | -0.373579 |
| 0.108453 | -0.00328464 |
| -0.219546 | -0.353415 |
| 0.0744453 | -0.460843 |
| -0.0664957 | -0.322419 |
| -0.285807 | -0.00693602 |
| -0.245026 | -0.139375 |
| 0.494443 | -0.358566 |
| -0.438267 | -0.145943 |
| -0.356567 | 0.0687614 |
| -0.505218 | 0.0249535 |
| -0.564685 | 0.124854 |

| F1 (16.40 %) | SAE (Obs) |
| --- | --- |
| -0.342441 | -0.223202 |
| 0.677209 | -0.530683 |
| -0.205416 | 0.173472 |
| 0.472728 | -0.00736532 |
| 0.0625958 | -0.373783 |
| -0.0182225 | -0.17092 |
| 0.316314 | -0.174122 |
| 0.555905 | -0.217346 |
| 0.161538 | -0.277176 |
| 0.433475 | -0.406936 |

| F1 (16.40 %) | TIF1 (Obs) |
| --- | --- |
| 0.390063 | -0.499308 |
| -0.0277319 | 0.106161 |
| 0.115087 | -0.437682 |
| -0.105157 | -0.171047 |
| -0.05883 | -0.202572 |
| 0.427279 | -0.304641 |
| -0.0561026 | 0.0098662 |
| -0.593869 | 0.172359 |
| 0.348272 | -0.295236 |
| 0.35768 | -0.423909 |
| 0.138925 | -0.468759 |
| -0.29493 | 0.0794928 |
| 0.362234 | -0.160464 |
| 0.45544 | -0.176665 |
| -0.619732 | 0.158583 |
| -0.0751794 | -0.377786 |
| -0.121281 | -0.094317 |
| 0.1528 | -0.306151 |
| 0.0243467 | -0.412552 |
| -0.121281 | -0.094317 |
| 0.266142 | -0.427318 |
| 0.0128778 | -0.33794 |
| 0.321065 | -0.282728 |
| -0.201542 | -0.246004 |
| -0.115169 | -0.0267636 |
| 0.582898 | -0.0883563 |
| 0.403222 | -0.169796 |
| 0.383759 | 0.572225 |
| -0.467506 | 0.0405776 |
| -0.24223 | -0.00374665 |
| -0.351631 | 0.181026 |
| 0.206979 | 0.125344 |
| -0.619732 | 0.158583 |
| -0.225881 | 0.171467 |
| -0.0390074 | -0.300071 |
| 0.174039 | -0.485952 |
| -0.220052 | -0.026318 |
| -0.143971 | 0.249451 |
| -0.520789 | 0.25519 |
| 0.272662 | -0.516492 |
| 0.261513 | -0.314733 |
| 0.256629 | -0.26588 |
| 0.233035 | -0.377641 |
| 0.158006 | -0.23534 |
| 0.652128 | 0.0670818 |
| 0.158006 | -0.23534 |
| 0.303906 | -0.0935951 |
| -0.0648069 | 0.0326728 |
| -0.00862641 | 0.151378 |
| -0.0461204 | 0.065211 |
| -0.0461204 | 0.065211 |
| -0.136199 | 0.0518971 |
| 0.552652 | -0.321921 |
| -0.0770989 | -0.455404 |
| 0.344686 | -0.226034 |
| -0.0484968 | -0.136277 |
| 0.453706 | -0.233476 |
| 0.387511 | -0.286968 |
| 0.456483 | -0.484397 |
| 0.0584796 | -0.200573 |
| -0.362906 | -0.225207 |
| -0.200929 | -0.368228 |
| -0.284106 | -0.158247 |
| -0.121281 | -0.094317 |
| 0.0150037 | -0.396769 |
| 0.0493063 | -0.39455 |
| -0.137567 | 0.0769884 |
| -0.0834234 | -0.383808 |
| -0.0842325 | -0.244006 |
| -0.0513154 | -0.297316 |
| 0.629662 | -0.384732 |
| 0.62976 | -0.0126947 |
| 0.856613 | 0.00763198 |
| 0.286459 | -0.128939 |
| 0.698805 | 0.142274 |
| 0.021814 | -0.367947 |
| 0.328337 | -0.401248 |
| 0.157103 | -0.231113 |
| 0.0490498 | -0.260225 |
| 0.471743 | -0.134244 |
| -0.39606 | -0.117662 |
| 0.544785 | -0.320221 |
| -0.126638 | 0.0478487 |
| -0.225881 | 0.171467 |
| 0.240279 | -0.441094 |

| F1 (16.40 %) | seronegative (Obs) |
| --- | --- |
| 0.402734 | -0.0645231 |
| -0.0997947 | 0.404139 |
| 0.102871 | 0.113493 |
| 0.180596 | -0.104311 |
| 0.349436 | 0.295233 |
| 0.824951 | -0.0150256 |
| -0.128104 | 0.187106 |

**B**

Symmetric plot (axes F1 and F2: 29.54 %)

| F1 (16.40 %) | Active variables |
| --- | --- |
| -0.101832 | 0.220119 |
| 0.201268 | -0.435058 |
| -0.116319 | -0.316421 |
| 0.638265 | 1.73626 |
| 0.176854 | -0.0514068 |
| -1.36605 | 0.397073 |
| 0.0684477 | 0.22594 |
| -0.140194 | -0.462769 |
| -0.0214798 | 0.0250633 |
| 0.521959 | -0.609037 |
| -0.0163895 | -0.00915055 |
| 0.575973 | 0.321576 |
| -0.371952 | -0.150686 |
| 0.662157 | 0.268253 |
| -0.680652 | -0.0103796 |
| 0.696987 | 0.0106287 |
| -0.0766405 | -0.190106 |
| 0.615861 | 1.52764 |
| -0.327567 | 0.508357 |
| 0.405835 | -0.629823 |
| -0.535891 | 0.167594 |
| 0.632905 | -0.197935 |
| -0.508139 | 0.140876 |
| 0.650052 | -0.18022 |
| -0.167056 | -0.046493 |
| 0.916665 | 0.255115 |
| -0.0307167 | -0.0346143 |
| 1.52355 | 1.71687 |
| -0.0760263 | -0.107699 |
| 1.29788 | 1.83857 |
| -0.212529 | -0.0278514 |
| 1.16618 | 0.152825 |
| -0.0115012 | 0.140821 |
| 0.0650728 | -0.79675 |
| -0.0744295 | -0.0354931 |
| 0.229291 | 0.109342 |
| -0.238217 | -0.30651 |
| 0.402942 | 0.518459 |
| 0.343888 | -0.0574169 |
| -0.99463 | 0.166067 |
| -0.168656 | 0.381959 |
| 0.280502 | -0.635259 |
| 0.548978 | -0.0967042 |
| -0.773797 | 0.136307 |
| -0.773797 | 0.136307 |
| 0.548978 | -0.0967042 |
| -0.195676 | 0.0561173 |
| 0.880544 | -0.252528 |
| -0.109009 | 0.0492233 |
| 0.912446 | -0.412017 |
| -0.307708 | -0.268985 |
| 0.854234 | 0.746733 |
| -0.167231 | -0.216199 |
| 1.19759 | 1.54827 |
| -0.476939 | 0.892174 |
| 0.272537 | -0.509814 |
| -0.291509 | 0.847577 |
| 0.187398 | -0.544871 |

| F1 (16.40 %) | Active observations |
| --- | --- |
| 0.306888 | 0.347464 |
| 0.530296 | -0.211747 |
| -0.776923 | 0.263554 |
| 0.213372 | -0.141922 |
| 0.390063 | -0.499308 |
| -0.659614 | 0.265552 |
| -0.0277319 | 0.106161 |
| 0.115087 | -0.437682 |
| 0.402734 | -0.0645231 |
| 0.433287 | 0.314678 |
| -0.105157 | -0.171047 |
| -0.05883 | -0.202572 |
| 0.499044 | 1.13173 |
| 0.427279 | -0.304641 |
| -0.308872 | 0.598366 |
| -0.0561026 | 0.0098662 |
| -0.593869 | 0.172359 |
| -0.0289415 | 0.557629 |
| -0.164857 | -0.0975064 |
| -0.776923 | 0.263554 |
| 0.478591 | 0.399756 |
| 0.348272 | -0.295236 |
| -0.0399664 | 0.00206994 |
| 0.35768 | -0.423909 |
| -0.0220519 | -0.489133 |
| 0.138925 | -0.468759 |
| -0.29493 | 0.0794928 |
| 0.362234 | -0.160464 |
| 0.625589 | 0.869732 |
| 0.45544 | -0.176665 |
| -0.619732 | 0.158583 |
| -0.0751794 | -0.377786 |
| -0.181207 | -0.27272 |
| -0.121281 | -0.094317 |
| 0.1528 | -0.306151 |
| 0.0243467 | -0.412552 |
| -0.419606 | -0.170381 |
| -0.121281 | -0.094317 |
| 0.405747 | -0.456454 |
| 0.266142 | -0.427318 |
| 0.0128778 | -0.33794 |
| 0.321065 | -0.282728 |
| 0.849558 | 0.871612 |
| -0.284131 | 0.373045 |
| -0.201542 | -0.246004 |
| -0.115169 | -0.0267636 |
| -0.494733 | 0.212602 |
| 0.582898 | -0.0883563 |
| -0.496257 | -0.218645 |
| -0.342441 | -0.223202 |
| 0.403222 | -0.169796 |
| 0.383759 | 0.572225 |
| 0.196703 | -0.444283 |
| -0.536384 | 0.0510696 |
| -0.0541311 | -0.251952 |
| -0.600857 | 0.0471394 |
| -0.776923 | 0.263554 |
| 0.288472 | 0.134353 |
| 0.29206 | 0.898668 |
| -0.467506 | 0.0405776 |
| -0.24223 | -0.00374665 |
| -0.776923 | 0.263554 |
| -0.663308 | 0.155394 |
| -0.776923 | 0.263554 |
| -0.351631 | 0.181026 |
| -0.244505 | -0.371417 |
| -0.776923 | 0.263554 |
| -0.0997947 | 0.404139 |
| 0.0761302 | -0.290317 |
| 0.206979 | 0.125344 |
| -0.619732 | 0.158583 |
| -0.225881 | 0.171467 |
| -0.0390074 | -0.300071 |
| -0.776923 | 0.263554 |
| 0.174039 | -0.485952 |
| -0.220052 | -0.026318 |
| -0.636617 | 0.0963496 |
| -0.143971 | 0.249451 |
| 0.216742 | 0.560076 |
| 0.247073 | 0.702971 |
| -0.520789 | 0.25519 |
| -0.641566 | 0.0817877 |
| -0.489432 | -0.146037 |
| -0.0685804 | -0.250526 |
| 0.272662 | -0.516492 |
| -0.434574 | -0.0357845 |
| 0.677209 | -0.530683 |
| 0.261513 | -0.314733 |
| 0.256629 | -0.26588 |
| 0.233035 | -0.377641 |
| -0.558708 | -0.11039 |
| -0.494733 | 0.212602 |
| 0.0724326 | -0.428244 |
| 0.501939 | -0.250182 |
| -0.307859 | -0.258936 |
| -0.226526 | -0.352231 |
| -0.558708 | -0.11039 |
| 0.119946 | 0.384865 |
| 0.158006 | -0.23534 |
| 0.54698 | 0.74589 |
| -0.776923 | 0.263554 |
| -0.205416 | 0.173472 |
| 0.102871 | 0.113493 |
| 0.402161 | -0.387252 |
| 0.0736494 | -0.10609 |
| 0.652128 | 0.0670818 |
| -0.118756 | -0.380975 |
| 0.158006 | -0.23534 |
| 0.303906 | -0.0935951 |
| -0.187436 | 0.429017 |
| 0.180596 | -0.104311 |
| -0.567569 | 0.188324 |
| -0.128468 | 0.592395 |
| -0.265763 | 0.278437 |
| 0.537455 | -0.242879 |
| -0.365507 | -0.190259 |
| 0.758328 | 1.23279 |
| 0.640923 | 0.715855 |
| -0.214077 | 0.0464884 |
| -0.0326861 | -0.374057 |
| -0.0648069 | 0.0326728 |
| 0.172272 | -0.136631 |
| -0.0708485 | 0.616662 |
| 0.472728 | -0.00736532 |
| 0.574504 | 0.700944 |
| 0.237618 | 0.0252241 |
| -0.00862641 | 0.151378 |
| -0.0461204 | 0.065211 |
| -0.262415 | -0.152939 |
| 0.921842 | 1.03966 |
| -0.622527 | 0.0229554 |
| -0.0461204 | 0.065211 |
| 0.048263 | -0.200933 |
| -0.663308 | 0.155394 |
| -0.136199 | 0.0518971 |
| -0.494733 | 0.212602 |
| -0.494733 | 0.212602 |
| 0.305334 | -0.240383 |
| -0.599488 | 0.022048 |
| -0.133172 | -0.134174 |
| -0.0513044 | -0.451285 |
| 0.0391409 | -0.0305708 |
| 0.609735 | -0.0867449 |
| 0.616255 | -0.175919 |
| 0.0191113 | 0.155573 |
| 0.552652 | -0.321921 |
| -0.367803 | -0.112244 |
| 0.7003 | -0.199816 |
| 0.101211 | -0.451294 |
| -0.122809 | -0.209251 |
| -0.0770989 | -0.455404 |
| -0.444788 | -0.0567688 |
| 0.344686 | -0.226034 |
| -0.622527 | 0.0229554 |
| 0.00629447 | 0.0531962 |
| -0.0484968 | -0.136277 |
| -0.274941 | -0.30333 |
| -0.0870246 | -0.143172 |
| -0.46413 | -0.159719 |
| -0.164857 | -0.0975064 |
| 0.453706 | -0.233476 |
| 0.387511 | -0.286968 |
| -0.0816805 | -0.307487 |
| 0.511417 | 0.105461 |
| 0.456483 | -0.484397 |
| 0.0584796 | -0.200573 |
| 0.0101296 | 0.495084 |
| -0.367803 | -0.112244 |
| -0.223591 | -0.372203 |
| 0.349436 | 0.295233 |
| 0.824951 | -0.0150256 |
| 0.617322 | -0.122824 |
| -0.295555 | -0.10251 |
| -0.362906 | -0.225207 |
| 0.171675 | -0.417595 |
| -0.200929 | -0.368228 |
| -0.284106 | -0.158247 |
| 0.474207 | 1.09149 |
| 0.0625958 | -0.373783 |
| 0.133204 | 0.758624 |
| 0.281166 | -0.184858 |
| -0.121281 | -0.094317 |
| -0.34423 | 0.475281 |
| 0.0150037 | -0.396769 |
| -0.624698 | 0.145549 |
| 0.0034689 | 0.427163 |
| 0.290796 | 0.891462 |
| -0.468042 | 0.153558 |
| 0.134572 | -0.208882 |
| 0.0493063 | -0.39455 |
| -0.677981 | 0.360161 |
| -0.0182225 | -0.17092 |
| -0.128104 | 0.187106 |
| 0.428167 | -0.137433 |
| -0.137567 | 0.0769884 |
| -0.776923 | 0.263554 |
| 0.409051 | 1.08378 |
| -0.0834234 | -0.383808 |
| -0.0842325 | -0.244006 |
| 0.27571 | 0.723454 |

| F1 (16.40 %) | F2 (13.14 %) |
| --- | --- |
| 0 | 0 |
| -0.101832 | 0.220119 |

| F1 (16.40 %) | F2 (13.14 %) |
| --- | --- |
| 0 | 0 |
| 0.201268 | -0.435058 |

| F1 (16.40 %) | F2 (13.14 %) |
| --- | --- |
| 0 | 0 |
| -0.116319 | -0.316421 |

| F1 (16.40 %) | F2 (13.14 %) |
| --- | --- |
| 0 | 0 |
| 0.638265 | 1.73626 |

| F1 (16.40 %) | F2 (13.14 %) |
| --- | --- |
| 0 | 0 |
| 0.176854 | -0.0514068 |

| F1 (16.40 %) | F2 (13.14 %) |
| --- | --- |
| 0 | 0 |
| -1.36605 | 0.397073 |

| F1 (16.40 %) | F2 (13.14 %) |
| --- | --- |
| 0 | 0 |
| 0.0684477 | 0.22594 |

| F1 (16.40 %) | F2 (13.14 %) |
| --- | --- |
| 0 | 0 |
| -0.140194 | -0.462769 |

| F1 (16.40 %) | F2 (13.14 %) |
| --- | --- |
| 0 | 0 |
| -0.0214798 | 0.0250633 |

| F1 (16.40 %) | F2 (13.14 %) |
| --- | --- |
| 0 | 0 |
| 0.521959 | -0.609037 |

| F1 (16.40 %) | F2 (13.14 %) |
| --- | --- |
| 0 | 0 |
| -0.0163895 | -0.00915055 |

| F1 (16.40 %) | F2 (13.14 %) |
| --- | --- |
| 0 | 0 |
| 0.575973 | 0.321576 |

| F1 (16.40 %) | F2 (13.14 %) |
| --- | --- |
| 0 | 0 |
| -0.371952 | -0.150686 |

| F1 (16.40 %) | F2 (13.14 %) |
| --- | --- |
| 0 | 0 |
| 0.662157 | 0.268253 |

| F1 (16.40 %) | F2 (13.14 %) |
| --- | --- |
| 0 | 0 |
| -0.680652 | -0.0103796 |

| F1 (16.40 %) | F2 (13.14 %) |
| --- | --- |
| 0 | 0 |
| 0.696987 | 0.0106287 |

| F1 (16.40 %) | F2 (13.14 %) |
| --- | --- |
| 0 | 0 |
| -0.0766405 | -0.190106 |

| F1 (16.40 %) | F2 (13.14 %) |
| --- | --- |
| 0 | 0 |
| 0.615861 | 1.52764 |

**C**

Dendrogram

|  | Dissimilarity |
| --- | --- |
| 1.96875 | 23.0909 |
| 1 | 23.0909 |
| 1 | 0 |
| 1 | 23.0909 |
| 2.9375 | 23.0909 |
| 2.9375 | 20.0078 |
| 2 | 20.0078 |
| 2 | 0 |
| 2 | 20.0078 |
| 3.875 | 20.0078 |
| 3.875 | 12.7174 |
| 3 | 12.7174 |
| 3 | 0 |
| 3 | 12.7174 |
| 4.75 | 12.7174 |
| 4.75 | 10.5063 |
| 4 | 10.5063 |
| 4 | 0 |
| 4 | 10.5063 |
| 5.5 | 10.5063 |
| 5.5 | 8.02848 |
| 5 | 8.02848 |
| 5 | 0 |
| 5 | 8.02848 |
| 6 | 8.02848 |
| 6 | 0 |
| 6 | 8.02848 |
| 5.5 | 8.02848 |
| 5.5 | 10.5063 |
| 4.75 | 10.5063 |
| 4.75 | 12.7174 |
| 3.875 | 12.7174 |
| 3.875 | 20.0078 |
| 2.9375 | 20.0078 |
| 2.9375 | 23.0909 |
| 1.96875 | 23.0909 |

|  | Dissimilarity |
| --- | --- |
| 1 | 7 |
| 2 | 7 |
| 3 | 7 |
| 4 | 7 |
| 5 | 7 |
| 6 | 7 |

**eFigure1. Histological features and clustering analysis**

A. Multiple correspondence analysis (MCA): observation plot. Anti-Mi-2 (magenta dots) and anti-MDA5 DM (green dots) were the most distinctive groups B. MCA: symmetric plot showing DMSA and closely associated pathology features. C. Agglomerate hierarchical clustering (AHC) dendrograms. Truncation: number of classes: 6 showing associations among different classes (see eTable 3)
