## Supplementary material for "Dermatomyositis: Muscle Pathology According to Antibody Subtypes": eFigure2

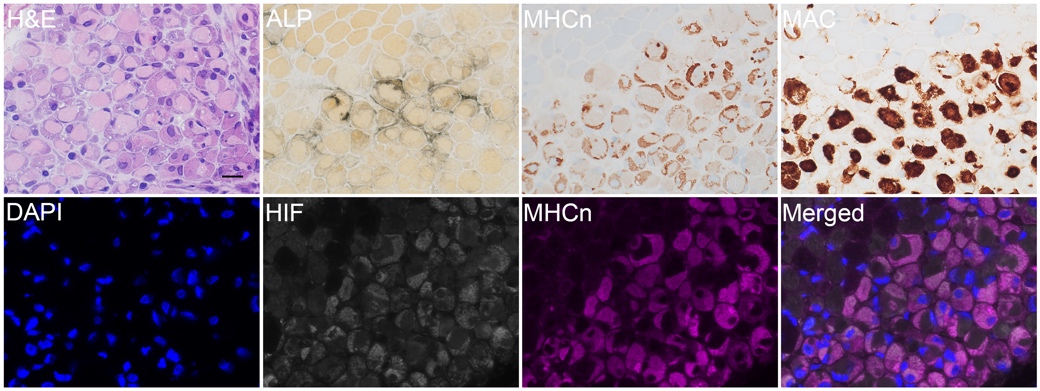

**eFigure 2 Central necrotic-peripheral regenerating fiber.** On H&E, the fibers present a homogenous pale eosinophilic necrotic center surrounded by crescent shaped basophilic regenerating portions at the periphery. The regenerating portions are highlighted by ALP, MHCn, and HIF The necrotic portions are highlighted by MAC. **Abbreviations:** H&E, hematoxylin and eosin; ALP, alkaline phosphatase; HIF, hypoxia inducible factor; MHCn, neonatal myosin; MAC, membrane attack complex.
